# Myonuclear apoptosis underlies diaphragm atrophy in mechanically ventilated ICU patients

**DOI:** 10.1101/2024.07.23.24310792

**Authors:** Wout J. Claassen, Marloes van den Berg, Zhong-Hua Shi, Rianne J. Baelde, Sylvia Bogaards, Luuk Bonis, Heleen Hakkeling, Arezou Bamyani, Gerben J. Schaaf, Albertus Beishuizen, Chris Dickhoff, Reinier A. Boon, Leo Heunks, Tyler J. Kirby, Coen A.C. Ottenheijm

## Abstract

**Rationale:** Mechanical ventilation plays an important role in critical illness-associated diaphragm weakness. Weakness contributes to difficult weaning and is associated with increased morbidity and mortality. Diaphragm weakness is caused by a combination of atrophy and dysfunction of myofibers, which are large syncytial cells maintained by a population of myonuclei. Each myonucleus provides gene transcripts to a finite fiber volume, termed the myonuclear domain. Changes in myonuclear number in myofibers undergoing atrophy has not been investigated in mechanically ventilated ICU patients. Myonuclear number is a determinant of transcriptional capacity, and therefore critical for muscle regeneration after atrophy.

**Objectives:** Our objective was to investigate if and how myonuclear number changes in the diaphragm of mechanically ventilated ICU patients and whether changes are associated with myofiber atrophy.

**Methods:** We used a combination of transcriptomics, immunohistochemistry, and confocal microscopy to study myonuclear alterations in diaphragm and quadriceps biopsies from mechanically ventilated ICU patients.

**Results:** Myonuclear number and myonuclear domain were reduced in patients with diaphragm myofiber atrophy. Intrinsic apoptotic pathway activation was identified as a mechanism underlying myonuclear removal in the diaphragm of mechanically ventilated ICU patients. Total transcriptional activity in myofibers decreased with myonuclear loss.

Furthermore, muscle stem cell number was reduced in the patients with diaphragm atrophy.

**Conclusion:** We identified myonuclear loss due to intrinsic apoptotic pathway activation as a potential mechanism underlying diaphragm atrophy in mechanically ventilated patients. This provides novel insights in diaphragm weakness of ICU patients. Targeted therapies may limit development of diaphragm weakness and improve weaning outcome.

## Introduction

Weakness of the diaphragm is a well-known consequence of critical illness and mechanical ventilation (MV) in patients admitted to the intensive care unit (ICU). It contributes to difficult weaning, which is associated with increased morbidity, mortality, and healthcare costs (1). Furthermore, it can lead to physical disability and long-term impairment in intensive care survivors (2). Diaphragm weakness is caused by a combination of atrophy and dysfunction of the remaining contractile material, leading to weakness of muscle cells (i.e., myofibers). In recent years, several mechanisms underlying contractile dysfunction have been identified (3–5), including changes in myosin conformation (6). The mechanisms underlying atrophy are not well understood. Increased proteolysis, resulting in a reduction of contractile material, may play a role (7, 8), but it is unclear whether other pathways contribute as well. Identifying the pathways that are involved in diaphragm myofiber atrophy will have important considerations for the development of therapies and for clinical practice, as diaphragm atrophy is associated with prolonged mechanical ventilation, longer ICU length of stay and increased risk of complications (9, 10).

Myofibers are large syncytial cells that are maintained by a population of post-mitotic myonuclei. Each myonucleus provides gene transcripts to a finite fiber volume, termed the myonuclear domain (11–14). During changes in muscle mass, myofibers’ nuclear number may change (15). In several pathological states resulting in myofiber atrophy, apoptosis regulates myonuclear number (16). In the diaphragm of ventilated rodents and brain-dead organ donors, myonuclear loss and upregulated apoptosis were identified to underlie diaphragm atrophy, but these pathways have not yet been investigated in ventilated ICU patients (17, 18). Furthermore, the brain-dead organ donors and rodents are very different from typical ICU patients, from perspective of ventilator mode, but also systemic inflammation and exposure to drugs; therefore, these models do not adequately reflect ICU patients (17). It is critical to investigate whether apoptosis and myonuclear loss occur in these patients as, during weaning, the diaphragm needs to regain mass and function for which nuclear number and their transcriptional activity are important (19, 20). Furthermore, loss of myonuclei may lead to longer-term functional impairment and weakness after hospital discharge, with have important consequences for patients including the risk for ICU readmission and persistent dyspnea (21). Finally, if myonuclear apoptosis plays a role in critical illness-associated diaphragm weakness, inhibiting the underlying mechanism may be a promising therapeutic venue (19, 20).

Thus, our objective was to study apoptotic pathways and myonuclear regulation in the diaphragm of mechanically ventilated ICU patients. Hence, we performed next-generation RNA-sequencing in diaphragm biopsies of MV ICU patients and immunofluorescence labeling of biopsy cryosections to investigate whether myonuclei undergo apoptosis.

Furthermore, we studied myonuclear number and morphology in myofibers isolated from diaphragm biopsies. Additionally, we obtained quadriceps biopsies of MV ICU patients to investigate whether our findings were specific to the diaphragm. Some of the results of these studies have been previously reported in conference abstracts and as a preprint (22, 23).

## Methods

For further details on the applied methods, see the online supplement.

### Patients, diaphragm biopsies

Diaphragm muscle biopsies were obtained from ICU patients receiving invasive mechanical ventilation (*ICU* patients, *n* = 24) and patients undergoing elective lung surgery for early-stage lung malignancy without critical illness (*Control* patients, *n = 10*). Exclusion criteria were chronic obstructive pulmonary disease (≥ GOLD stage III), congestive heart failure, neuromuscular diseases, chronic metabolic disorders, pulmonary hypertension, chronic use of corticosteroids (> 7.5mg/day for at least 3 months), and more than 10% weight loss within the last 6 months. The exclusion criteria were similar for all experimental groups. Patients’ characteristics are presented in Table 1. The biopsy protocol was approved by the institutional review board at Amsterdam UMC (location VUmc), the Netherlands. Patients were recruited in Amsterdam UMC, the Netherlands Cancer Institute-Antoni van Leeuwenhoek Hospital (both in Amsterdam, the Netherlands), and Medisch Spectrum Twente (Enschede, the Netherlands). Written informed consent was obtained from the patients or their legal representative.

**Table 1.** Clinical characteristics of patients.

| Characteristic | Control<br>(n = 10) | ICU<br>Patients<br>(n=24) | ICU A+<br>(n =14 ) | ICU A-<br>(n=10) | P-value |
| --- | --- | --- | --- | --- | --- |
| Age (years) | 67 [60-72] | 66 [50-73] | 65 [55-71] | 60 [49-69] | 0.698 |
| Male (%) | 7 (70) | 15 (63) | 7(50) | 8 (80) |  |
| BMI (Kg/m<sup>2</sup>) | 28 [24-32] | 25 [21-28] | 25 [21-28] | 26 [21-28] | 0.281 |
| APACHE-3 | - | 73 [47-111] | 78 [53-149] | 75[67-89] | 0.762 |
| Ventilation (hours) | 1.5 [0.9-2.0] | 66 [44-193] | 63 [45-121] * | 90 [18-223] * | <0.001 |
| **Controlled ventilation** (hours) | 2.0 [1-2.3] | 64 [32-127] * | 64 [34-132]* | 55 [16-140]* | <0.001 |
| Myofiber CSA (μm<sup>2</sup>) | 2469[1708-2973] | 1736 [1250-3035] | 1341 [1007-1667] * | 3159 [2798-4294] * | <0.001 |
| **Medical history, n (%)** |  |  |  |  |  |
| Smoking | 3 (30) | 13 (54) | 10(71) | 3 (30) | 0.072 |
| COPD ≤ G2 | 3 (30) | 3 (13) | 2 (14) | 1 (10) | 0.506 |
| Other lung disease | 2 (20) | 1 (4) | 1 (7) | 0 (0) | 0.259 |
| Cardiac | 1 (10) | 3 (13) | 2 (14) | 1 (10) | 0.996 |
| Arterial vascular disease | 2 (20) | 14 (58)* | 12 (86) * | 2 (20) | 0.010 |
| Hypertension | 3 (30) | 9 (38) | 6 (43) | 3 (30) | 0.860 |
| CKD | 1 (10) | 2 (8) | 2 (14) | 0 (0) | 0.535 |
| T2DM | 0 (0) | 2 (8) | 0 (0) | 2 (20) | 0.179 |
| Hypothyroidism | 1 (10) | 2 (8) | 2 (14) | 0 (0) | 0.716 |
| Malignancy lung | 7 (70) | 1 (4) * | 0 (0) * | 1 (10) * | <0.001 |
| Malignancy other | 2 (20) | 2 (1) | 2 (1) | 0 (0) | 0.466 |
| **Medication** |  |  |  |  |  |
| Steroids | - | 18 (75) | 11 (79) | 9 (90) | 0.715 |
| Neuromuscular blockers | - | 12 (50) | 8 (57) | 4 (40) | 0.433 |
| Vasopressors | - | 19 (79) | 10 71) | 9 (90) | 0.495 |

Data displayed as Median [IQR], p-values calculated with Kruskal-Wallis or chi-squared tests BMI: Body Mass Index, MVhr: duration of mechanical ventilation before biopsy *BMI = Body Mass Index* APACHE = Acute Physiology And Chronic Health Evaluation, CSA = Cross-Sectional Area, COPD = Chronic Obstructive Pulmonary Disease, CKD = Chronic Kidney Disease, T2DM = Type 2 Diabetes Mellitus, CID = Chronic Inflammatory Disease, P/F-ratio = arterial partial pressure of oxygen (PaO<sub>2</sub>) divided by the inspired oxygen concentration (FiO<sub>2</sub>), A-a gradient = alveolar-arterial gradient. Data shown as Median [IQR]. P-values of continuous data calculated with one-way analysis of variance or Kruskal-Wallis test, depending on distribution of the data. P-values of categorical data calculated with Chi-squared test. \* indicates a significant difference with the control group calculated with post-hoc tests.

**Table 2.** Characteristics of individual patients of quadriceps studies.

| Patient | Age (y) | Sex | BMI (kg/m<sup>2</sup>) | ICU diagnosis | APACHE-4 score | Duration of MV (h) |
| --- | --- | --- | --- | --- | --- | --- |
| **1** | 56-60 | M | 26 | Trauma | 139 | 33.3 |
| **2** | 80-85 | M | 25 | Trauma | 94 | 44.3 |
| **5** | 30-35 | F | 27 | Trauma | 76 | 50.5 |
| **6** | 30-35 | M | 22 | Trauma | 76 | 50.8 |
| **8** | 80-85 | M | 23 | Sepsis | 130 | 28.3 |
| **9** | 60-65 | M | 25 | Trauma | 94 | 17 |
| **10** | 20-25 | M | 24 | Trauma | 63 | 79 |
| **11** | 70-75 | M | 29 | Trauma | 40 | 5 |
| **14** | 70-75 | M | 28 | Resp. failure | 97 | 49 |
| **15** | 66-70 | M | 20 | Resp. failure | 104 | 4 |
| **C010** | 20-25 | M | 23 | - | - | - |
| **C011** | 20-25 | M | 22 | - | - | - |
| **C-09** | 56-60 | M | 26 | - | - | - |
| **C-10** | 50-55 | F | 23 | - | - | - |
| **C-11** | 56-60 | M | 32 | - | - | - |

### Patients, quadriceps biopsies

Quadriceps biopsies of ICU patients (*n* = 10) were obtained in the context of a separate study that has been filed in the Clinical Trial Register under #NCT03231540 and was approved by the Medical Ethical Committee of VU Medical Center, Amsterdam, the Netherlands. Informed consent was obtained from the patient or a legal representative. Patients ≥18 years that had an expected ventilation duration of ≥72 hours, were expected to tolerate enteral nutrition ≥72 hours, and had a Sequential Organ Failure Assessment (SOFA) score ≥6, were considered eligible for inclusion. Exclusion criteria were contra-indications to enteral nutrition, short bowel syndrome, type C liver cirrhosis or acute liver failure, dependency on renal replacement therapy, requiring another specific enteral nutrition formula for medical reasons, BMI >35 kg/m2, extensive treatment limitations, disseminated malignancy, hematological malignancy, primary neuromuscular pathology, chronic use of corticosteroids for >7 days before ICU admission or contra-indications for muscle biopsy such as the need for continuous systemic anticoagulation, INR >1.3 or thrombocytes <100 × 10^3^ / mm^3^.

### Single myofiber microscopy

Single myofibers were manually isolated from the biopsies. Myonuclear number, myofiber volume, myonuclear morphology, and RNA-polymerase-II Ser5 fluorescence were determined using immunofluorescence labeling in combination with confocal microscopy.

### RNA-sequencing

RNA was extracted using a commercially available kit and sequenced using a NextSeq500 (Illumina).

### Immunohistochemistry

Serial cryosections were cut from the biopsies and stained to study myonuclear number, myonuclear domain, markers for apoptosis, and satellite cell content.

### Statistical analysis

Normality of the distribution of the studied variables was assessed visually on normal probability plots. Log transformation was performed if necessary. To compare the difference between ICU patients and control patients, student’s t-test or Mann-Whitney-U was applied for non-repeat measurements; and linear mixed model with patients as the random factor was applied for measurements involving technical replicates in all human samples. For linear mixed models, Greenhouse-Geisser correction was applied to adjust for potential lack of sphericity. For the comparison of three groups or more, one-way analysis of variance (ANOVA) or Kruskal-Wallis tests were performed with Tukey’s or Dunn’s post-hoc tests, depending on the distribution of the data. We used a two-sided significance level of 5% for all analyses. Unless otherwise noted, data are expressed as mean (± standard error), median [interquartile range], or frequencies (percentage), as appropriate.

## Results

Diaphragm biopsies were collected from 24 mechanically ventilated ICU patients who underwent laparotomy or thoracotomy for a clinical indication. The biopsies were collected from the left anterolateral part of the zone of apposition of the diaphragm. In Table 1 we summarize the clinical parameters. The biopsies were compared to the biopsies of 10 patients who underwent elective thoracic surgery for a small, primary pulmonary nodule. The groups were matched for age, body mass index, and sex (Table 1). Due to the limited size of the biopsies, not every biopsy was used in every experiment. Table S1 details which biopsies were used for each experiment.

### Transcriptomics reveals upregulation of apoptotic pathways in the diaphragm

To investigate the mechanisms underlying diaphragm weakness and atrophy, we performed next-generation RNA-sequencing of whole tissue samples cut from the biopsies. Table S2 shows the clinical characteristics of the patients included in this experiment. Principal component analysis (PCA) revealed clear clustering in the control patients, while the ICU group showed a more variable clustering pattern (Fig. 1A). We identified 2977 differentially expressed genes (1741 upregulated in the ICU group and 1236 downregulated (Fig. 1B) between control and ICU patients. A heatmap of the top 50 differentially expressed genes was generated (Fig. 1C). Next, we performed Panther pathway enrichment analysis of significant (FDR < 0.2, dashed line represents *p*-value of 0.05) differentially expressed genes (Fig. 1D). Consistent with our hypothesis of increased apoptosis in ICU patients, the top upregulated pathways in the ICU group were the p53, apoptosis, and integrin signaling pathways. Thus, in mechanically ventilated ICU patients, the p53 and apoptosis pathways are upregulated (Fig. 1E, S1A), providing a potential mechanism underlying weakness and atrophy.

**Figure 1.**
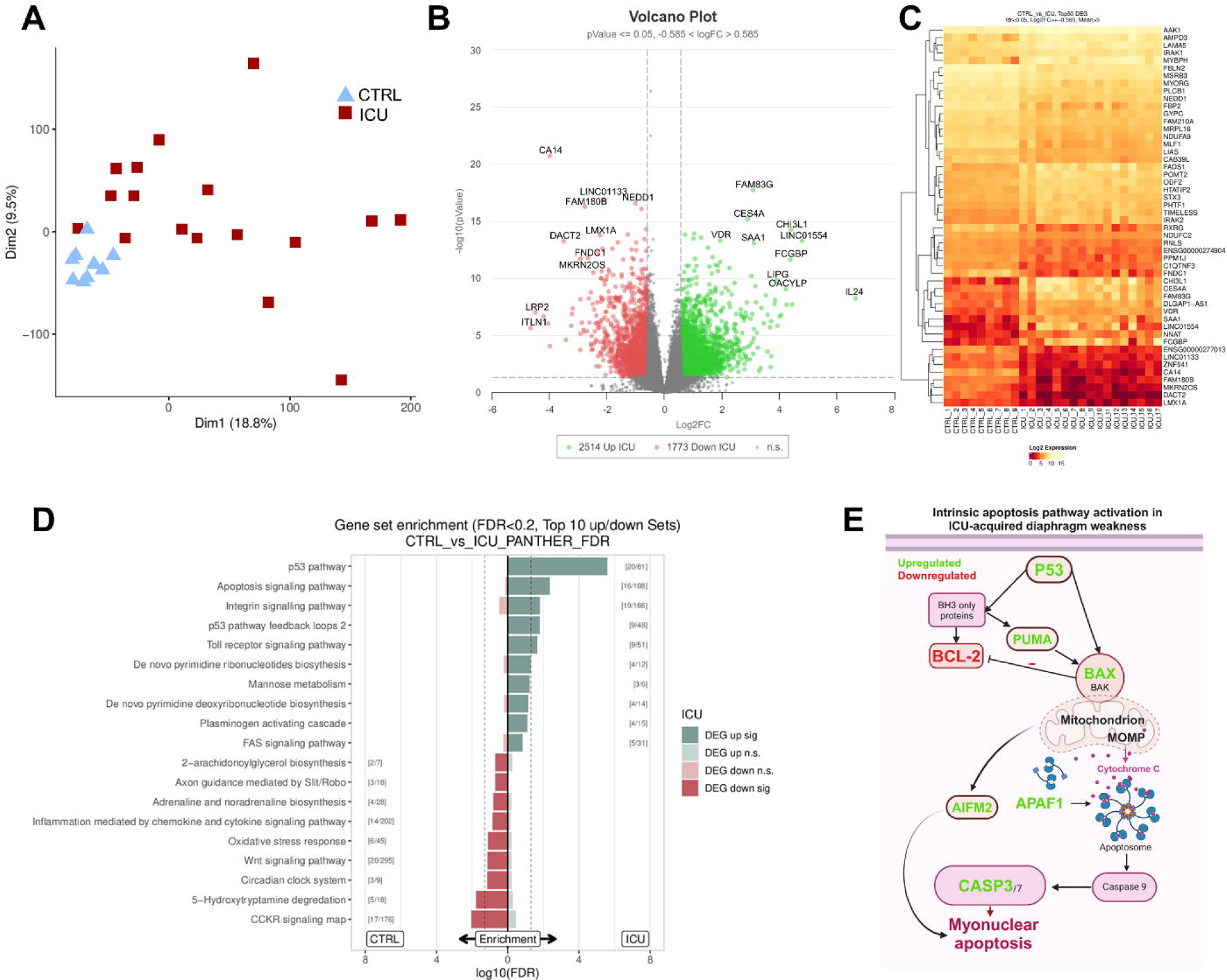
RNA sequencing of the diaphragm of mechanically ventilated ICU patients. **A**: Principal component analysis of sequencing results. Note the clustering of the samples within the CTRL group while this clustering is absent in the ICU groups. This may be due to heterogeneity of patient characteristics within both ICU groups. CTRL *N* = 8; ICU *N* = 17. **B:** Volcano plot and **C:** top 50 DEG heatmap. Top 50 most significantly differentially expressed DEGs for each contrast (sorted by smallest adjusted *p*-value). 1714 genes were significantly upregulated and 1236 genes were significantly downregulated in the ICU group. Genes with a significance level *p*<0.05 and a fold change of >1.5 were deemed differentially expressed. CTRL *N* = 8; ICU *N* = 17. **D:** PANTHER gene set enrichment analysis. Top10 gene sets or pathways enriched for up- or down-regulated genes of one database (dashed line: *p*-value = 0.05). Only shows pathways that are not significant for both directions (up/down) at the same time to identify on/off situations. Significant gene set enrichment is defined by the false discovery rate. CTRL *N* = 8; ICU *N* = 17. **E:** Schematic of the differentially expressed genes that are involved in the intrinsic apoptotic pathway. Genes in green are significantly upregulated and genes in red are significantly downregulated in ICU patients.

### Caspase-3-mediated apoptosis of myonuclei

As bulk-tissue RNA-sequencing revealed marked upregulation of the apoptotic pathway, next we performed analyses on muscle cross-sections to allow for the identification of different cell types that may be undergoing apoptosis. We used multiple markers to distinguish myonuclei from non-myonuclei (e.g. pericentreolar-material-1 (PCM1) (24) and dystrophin/laminin immunolabeling), as previously it was suggested that much of the apoptosis that occurs during muscle atrophy can be attributed to the non-myonuclear cell pool (24). Tables S3 and S4 show the characteristics of the patients included in these experiments. The percentage of myonuclei undergoing apoptosis was determined using terminal deoxynucleotidyl transferase dUTP nick end labeling (TUNEL) of double-stranded DNA breaks (Fig. 2A). Representative images of TUNEL labeling of a DNase-I treated diaphragm section (positive control) are shown in Fig. S2. The mean TUNEL-index (number of TUNEL-positive myonuclei divided by the total number of myonuclei) in the ICU group was almost double that of the control group (Fig. 2B). There was no difference in TUNEL-index for other cell types (Fig. S3A). Furthermore, we used an antibody for cleaved (i.e. activated) caspase-3 to investigate the role of caspase-3 mediated nuclear apoptosis (Fig. 2C) (25).

**Figure 2.**
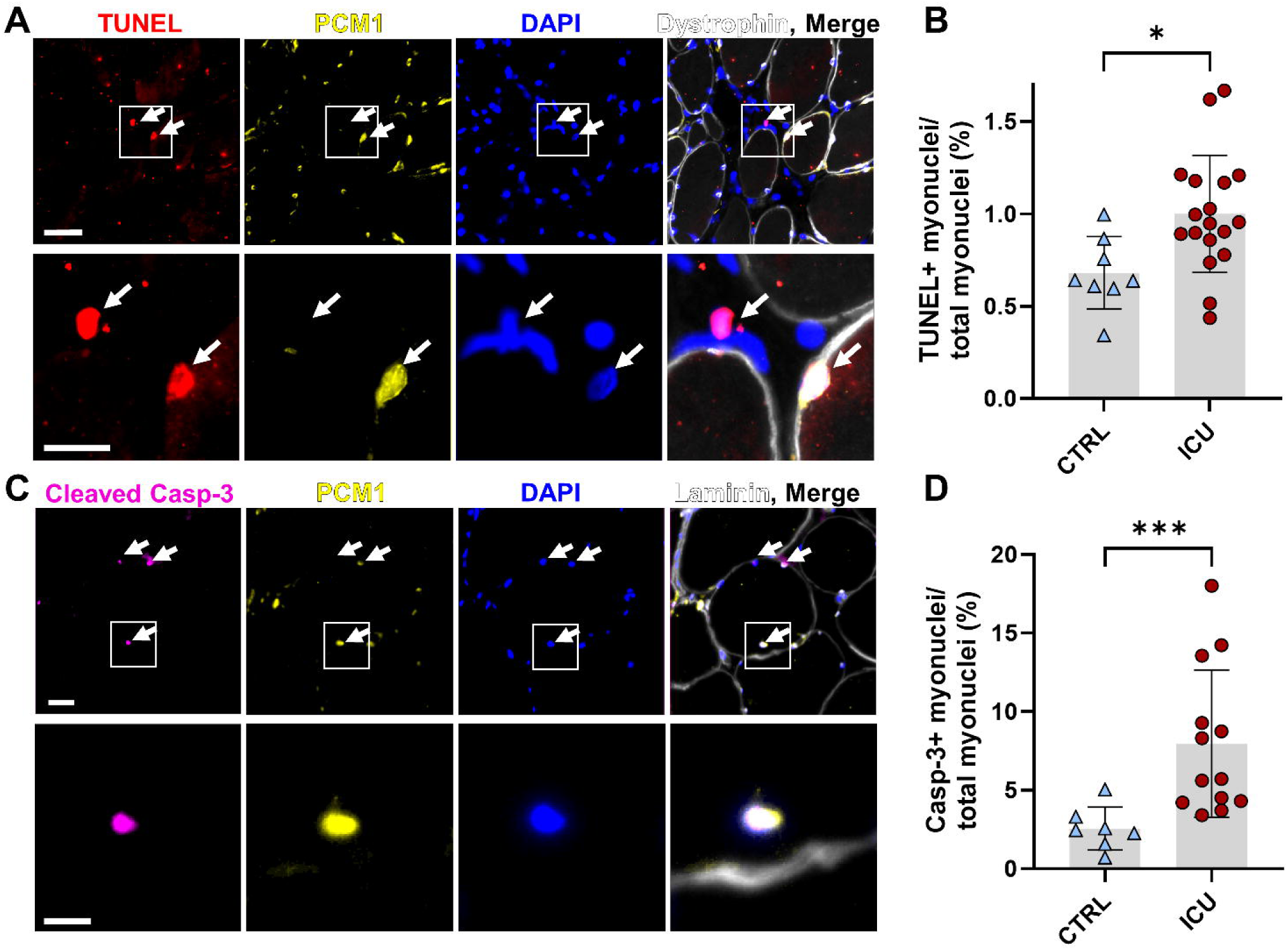
Caspase-3 mediated apoptosis as a mechanism underlying myonuclear loss in the diaphragm of ICU patients. **A**: Representative images of diaphragm muscle cross-sections stained with TUNEL assay, PCM1 antibody, Dystrophin antibody, and DAPI. Nuclei with a TUNEL-positive signal were designated as apoptotic myonuclei when they were PCM1 positive and were located within the dystrophin barrier. Note the difference in PCM1 immunoreactivity of the TUNEL+ non-myonucleus (left white arrow) and myonucleus (right white arrow). Scale bar top row is 50 µm, scale bar bottom row is 20 µm. **B:** Quantification of TUNEL index, calculated as the percentage of TUNEL-positive myonuclei. Total myonuclear count was determined by counting PCM1-positive nuclei. The grey bar represents the mean ± standard deviation within the groups of patients. Each colored symbol represents the value of a single patient. CTRL *N* = 8; ICU *N* = 18. Significance level was calculated using an unpaired *t*-test. **C:** Representative images of diaphragm muscle cross-sections stained with Cleaved caspase-3 antibody, PCM1 antibody, Laminin antibody and DAPI. Nuclei with a Cleaved Caspase-3 positive signal were designated as apoptotic myonuclei when they were PCM1 positive and were located within the laminin barrier. Scale bar top row = 50µm, scale bar bottom row = 20 µm. **D:** Quantification of activated caspase-3 index, calculated as the percentage of activated caspase-3-positive myonuclei. Total myonuclear count was determined by counting PCM1-positive nuclei. The grey bar represents the mean ± standard deviation within the groups of patients. Each colored symbol represents the value of a single patient. CTRL *N* = 7; ICU *N* = 14. Significance level was calculated using a Mann-Whitney-U test. ICU A+ = ICU group with atrophy, ICU A− = ICU group without atrophy, CTRL = Control group, * = *p*<0.05, ** = *p*<0.01 Figure 3. Reduced number of myonuclei in atrophic diaphragm fibers of critically ill patients.

The mean activated caspase-3 index (number of cleaved caspase-3-positive myonuclei divided by the total number of myonuclei) was ∼3-fold higher in the ICU group (Fig. 2D). Caspase-3-index for other cell types was also higher in the ICU group (Fig. S3B).

Next, to investigate whether our findings are specific to the diaphragm, we obtained quadriceps muscle biopsies (*n = 10*) and compared them to healthy controls (*n = 5*). Tables 2 and S5 show the clinical characteristics of these groups. Note that the duration of mechanical ventilation and disease severity is similar to that of the groups from which diaphragm biopsies were obtained. The percentage of myonuclei undergoing apoptosis was determined using the same activated caspase-3 immunofluorescence protocol used for the diaphragm biopsies (Fig. S4A). In the ICU group, the activated caspase-3 index of both myonuclei and other cell types was almost double that of the control group, but still more than two-fold lower than in the diaphragm (Fig. S4B, C, S5). The mean myofiber CSA was similar in the ICU and control groups (Fig. S4D), suggesting that in the quadriceps of MV ICU patients, caspase-3 activation occurs before the onset of atrophy.

Thus, both our transcriptomic and immunofluorescence data support a mechanism where caspase-3-mediated apoptosis is increased during critical illness-associated diaphragm weakness and is increased in quadriceps muscle before the onset of atrophy.

### Decreased myonuclear number and myonuclear domain in atrophic diaphragm fibers

To investigate whether the upregulation of apoptosis in myonuclei resulted in a reduced myonuclear number, we counted myonuclei in 8-12 myofibers per biopsy that were randomly selected and manually isolated. This method allows for a precise determination of myonuclear number per myofiber volume. Myonuclear domain was determined by dividing myofiber volume by myonuclear number. Immunofluorescent staining of myonuclear marker PCM1 revealed that the proportion of PCM1 positive myonuclei within the isolated myofibers (*n* = 3 control and *n* = 3 ICU, Table S6) was >98% in both groups, showing that the number of non-myonuclei included in the preparations did not impact nuclear number (Fig. S6). Fast and slow-twitch fibers were identified by immunoreactivity for fast-twitch myosin heavy chain isoform and data were segregated according to fiber type.

Myofiber volume and myonuclear domain size were lower in the ICU group, but nuclear number was not significantly lower (Fig. S7). We hypothesized that we did not observe a difference in myonuclear number due to the presence of patients in the ICU group with a myofiber volume similar to that of the controls. Therefore, to investigate whether myonuclear number was reduced specifically in ICU patients with atrophy, we separated the ICU group into two groups: one with atrophy (ICU A+) and one without atrophy (ICU A−). To this end, we used myofiber CSA data derived from cryosections (mean size of ∼450 myofibers). For the ICU A−-group, the lower limit of the CSA was set at the median CSA of the control group (CSA of 2500 µm^2^). To ensure minimal overlap between the groups, the upper limit of the ICU A+ group was set at a CSA of 2000 µm^2^. Clinical characteristics of all groups are shown in Table 1.

As expected, the volume of the isolated myofibers in the ICU A+ group was ∼50% smaller compared to the ICU A− and control groups, and this difference was similar for both fiber types (Fig. 3C). In the ICU A+ group, myonuclear number was reduced by about 35% in both fast and slow myofibers when compared to the ICU A− and the control group (Fig. 3D). To verify whether our findings in single myofibers were representative of a larger sample, we validated these findings by analyzing the number of myonuclei per myofiber in diaphragm muscle cross-sections with an average size of 498 myofibers per section. Myonuclei were identified using PCM1 labeling in combination with localization within the dystrophin barrier (note that we did not stain for myofiber type). In muscle cross-sections, average myonuclear counts per fiber were reduced by about 39% in the ICU A+ group compared to the control group (Fig. S8B), which was consistent with the results from the isolated myofibers (Fig. 3D). Next, we counted the myonuclei present in the quadriceps cross-sections, and found that there was no difference in the number of myonuclei per myofiber between the control and ICU patients (Fig. S8C). In the isolated myofibers, myonuclear domain size was smaller in the ICU A+ group when compared to the control group, but not when compared to the ICU A− group (Fig. 3E). When we examined the relationship between myofiber CSA and myonuclear number, we found a comparable positive correlation in all three groups (Fig.

**Figure 3.**
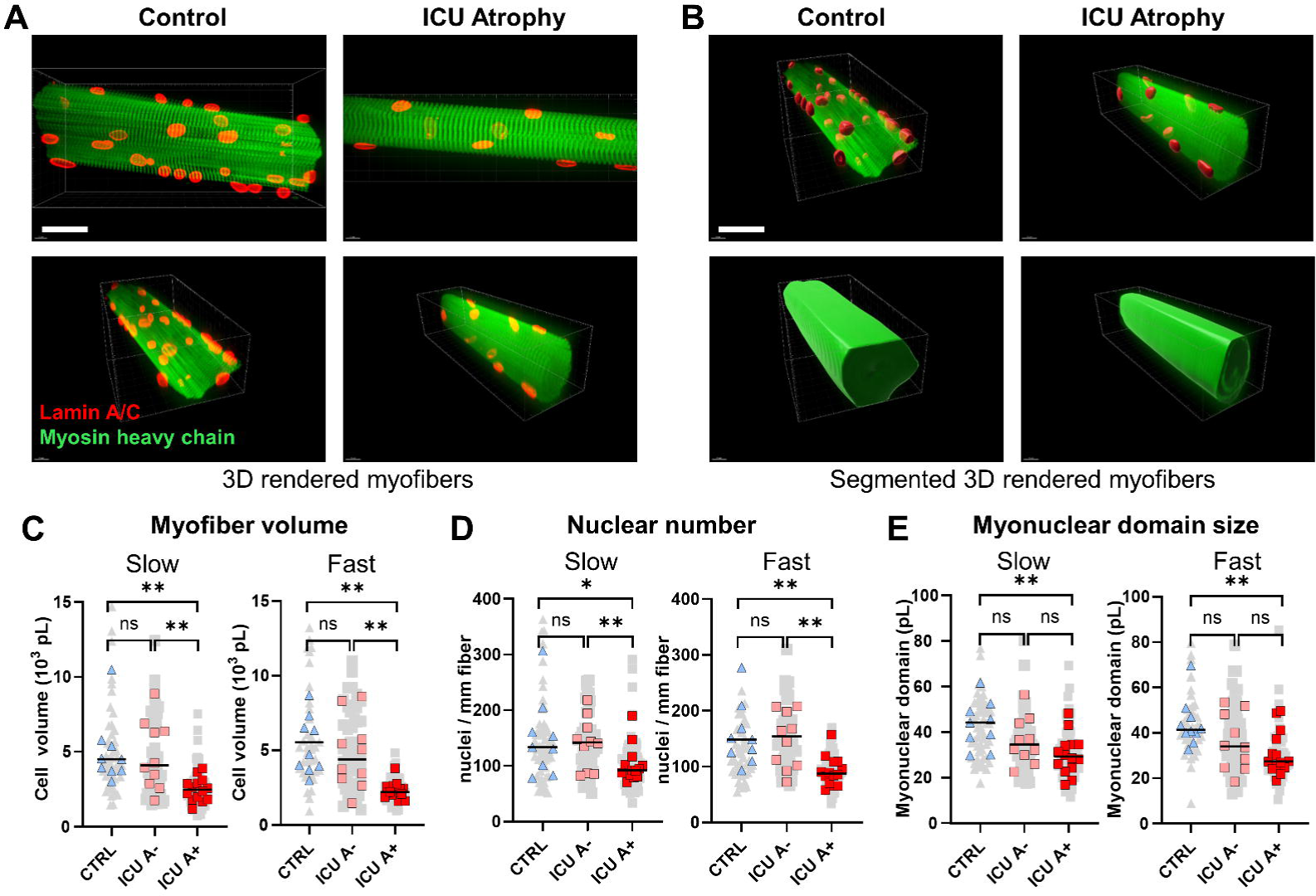
Reduced number of myonuclei in atrophic diaphragm fibers of critically ill patients. **A**: Representative images of single muscle fibers from control (left) and ICU (right) patients. Myofibers are immunofluorescently labeled for lamin A/C (red) and myosin heavy chain (green) Scale bar = 60 µm **B:** Representative images of 3D rendered single muscle fibers, with segmentation in 3D. Scale bar = 60 µm. **C:** Quantification of myofiber volume, calculated as volume per mm fiber was normalized to a sarcomere length of 2.5 µm. Every grey dot represents the value of a single muscle fiber and the colored symbols represent the mean values of a single patient. Slow-twitch fibers: ICU A+ *N =* 14, *n* = 84; *N =* 10, *n =* 50; CTRL *N =* 10, *n =* 48. Fast-twitch fibers: ICU A+ *N =* 14, *n =* 56; ICU A− *N =* 10, *n =* 48; CTRL *N =* 10, *n =* 46. **D:** Quantification of myonuclear number. Every grey dot represents the value of a single muscle fiber and the colored symbols represent the mean values of a single patient. Black bars indicate the median of the whole group. Slow-twitch fibers: ICU A+ *N =* 14, *n =* 84; ICU A− *N =* 10, *n =* 50; CTRL *N =* 10, *n =* 48. Fast-twitch fibers: ICU A+ *N =* 14, *n =* 56; ICU A− *N =* 10, *n =* 48; CTRL *N =* 10, *n =* 46. **E** Quantification of myonuclear domain size. Every grey dot represents the value of a single muscle fiber and the colored symbols represent the mean values of a single patient. Significance levels were calculated using linear mixed models with the patients as the random factor. Black bars indicate the median of the whole group. Slow-twitch fibers: ICU A *N =* 14, *n =* 84; ICU A− *N =* 10, *n =* 50; CTRL *N =* 10, *n =* 48. Fast-twitch fibers: ICU A+ *N =* 14, *n =* 56; ICU A− N = 10, *n =* 48; CTRL *N =* 10, *n =* 46. ICU A+ = ICU group with atrophy; ICU A− = ICU group without atrophy; CTRL = Control group. * denotes *p* < 0.05; ** denotes *p* < 0.01. *N =* number of patients, *n =* number of analyzed myofibers.

S9A). Myonuclear domain size and myofiber CSA showed a similar positive correlation, but the elevation of the regression line in the ICU A+ group was significantly lower, indicating smaller myonuclear domains for the same CSA. The slope of the regression line of the fibers isolated from the biopsies of ICU A− patients had a significantly steeper relationship between myonuclear domain and cross-sectional area compared to the ICU A− group (Fig. S9B). In conclusion, myonuclear number is reduced in myofibers in the atrophic diaphragm of mechanically ventilated ICU patients, and myonuclear domain is smaller, indicating relatively more atrophy than myonuclear loss. Furthermore, the reduction in myonuclear number was absent in the diaphragm of similar ICU patients without established atrophy.

Thus, atrophic diaphragm myofibers have a reduced number of myonuclei. In the diaphragm, loss of myonuclei appears to commence before the onset of atrophy, and before changes in myonuclear number in limb muscle. This may have profound implications for weaning and recovery after ICU and hospital discharge, as it has been proposed that myofibers depend on their myonuclear content for recovery after atrophy (26).

### Changes in myonuclear morphology

In addition to myonuclear number, myonuclear size is known to scale with myofiber size (27). Based on the changes we observed in myonuclear number, we sought to study changes in myonuclear morphology by segmenting every 3D-rendered nucleus present within the Z-stacks of the myofibers (Fig. 4A). Myonuclear volume was reduced in ICU A+ patients (Fig. 4B), and the cumulative fluorescence intensity of the nuclear lamina (lamin A/C) within each nucleus was increased (Fig. 4C). An increased total fluorescent intensity of the nuclear lamina may indicate increased nuclear wrinkling or increased thickness of the nuclear lamina. To further study this, we determined the wrinkling index of each nucleus by measuring the standard deviation of the fluorescence intensity within each segmented nucleus in maximum intensity Z-projections of the Z-stacks (Fig. 4D) (28). Nuclear wrinkling was increased in the ICU A+ and A− groups, when compared to the control group (Fig. 4E). Mean ellipticity or sphericity, two shape descriptors of sphere-like objects, were not different between the groups (Fig. S10 A-B). Thus, nuclear volume was lower in the ICU A+ group and myonuclear wrinkling was increased in in both ICU A+ and ICU A− patients.

**Figure 4.**
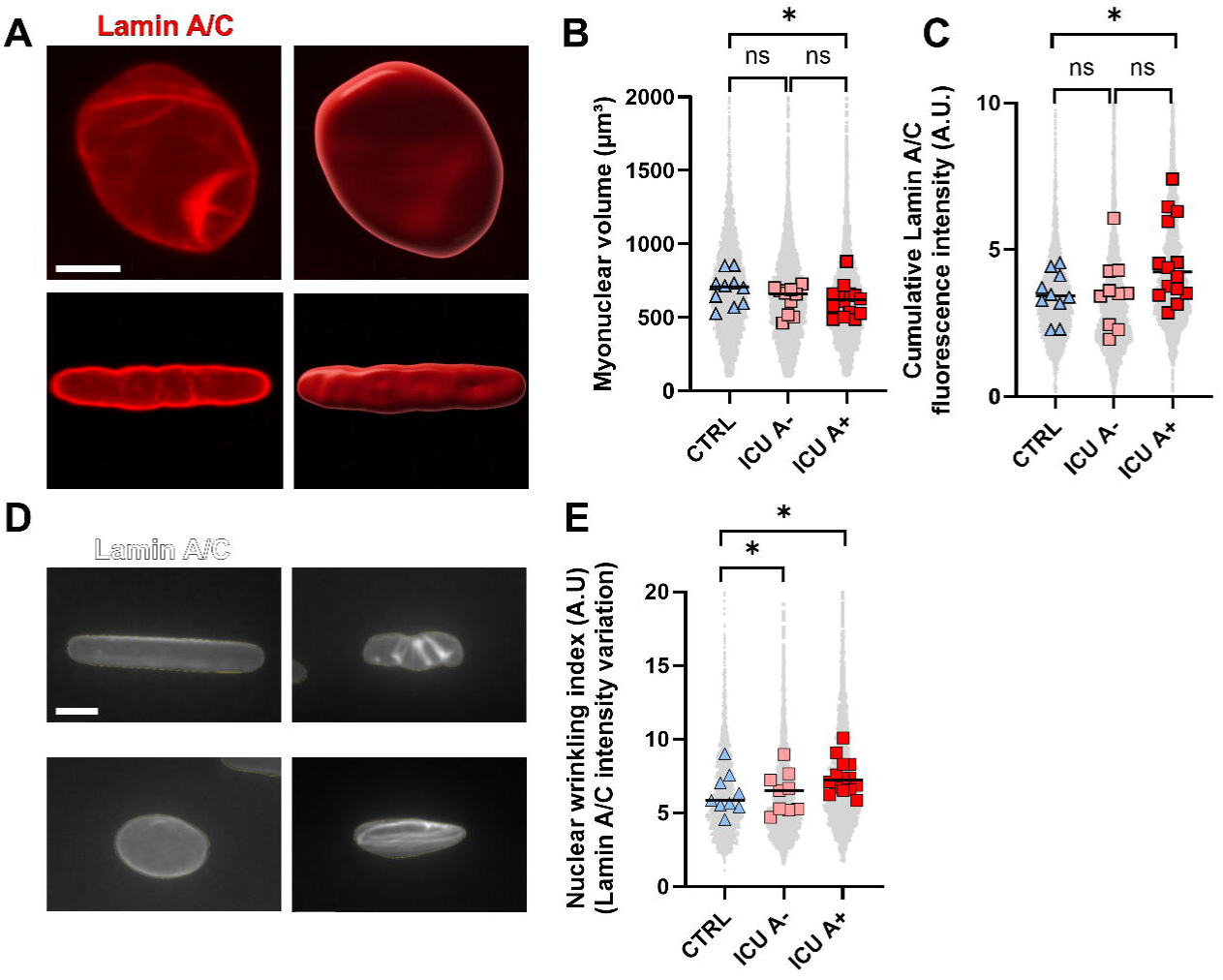
Altered myonuclear morphology in diaphragm myofibers of ICU patients. **A**: Representative images of unsegmented (left) and segmented (right) 3D-rendered myonuclei within mounted single muscle fibers that were immunofluorescently labeled for lamin A/C. Note the difference in ellipticity and sphericity between the top and the bottom myonucleus. Scale bar = 5 µm **B** Quantification of myonuclear volume. Each grey dot represents the value of a single nucleus. Each colored symbol represents the mean value of a single patient. The black bar represents the mean value within the groups of patients. Significance level was calculated using linear mixed models with the patients as the random factor. CTRL *N =* 10 patients, *n* = 6270 nuclei; ICU A+ *N =* 14 patients; *n* = 5463 nuclei; ICU A− *N = 10* patients; *n* = 6330 nuclei. **C** Quantification of lamin A/C fluorescence intensity within each nucleus. Each grey dot represents the value of a single nucleus. Each colored symbol represents the mean value of a single patient. The black bar represents the mean value within the groups of patients. Significance level was calculated using linear mixed models with the patients as the random factor. CTRL *N =* 10 patients, *n* = 6270 nuclei; ICU A+ *N =* 14 patients, *n* = 5463; ICU A− *N =* 10 patients, *n* = 6330 nuclei. . **D:** Representative images of maximum intensity Z-projections of Z-stacks of myonuclei. Note the difference between the nuclei with a smooth nuclear lamina (left) and a wrinkled nuclear lamina (right). Scale bar = 5 µm. **E:** Quantification of the wrinkling index for each nucleus. Wrinkling index was calculated as the standard deviation of the fluorescence intensity within the segmented nucleus. Each grey dot represents the value of a single nucleus. Each colored symbol represents the mean value of a single patient. The black bar represents the mean value within the groups of patients. Significance level was calculated using linear mixed models with the patients as the random factor. CTRL *N =* 10 patients, *n* = 1983 nuclei; ICU A+ *N =* 14 patients, *n* = 2601 nuclei; ICU A− *N =* 10 patients, *n* = 2442 nuclei. ICU A+ = ICU group with atrophy; ICU A− = ICU group without atrophy; CTRL = Control group. * denotes *p* < 0.05; ** denotes *p* < 0.01.

**Figure 5.**
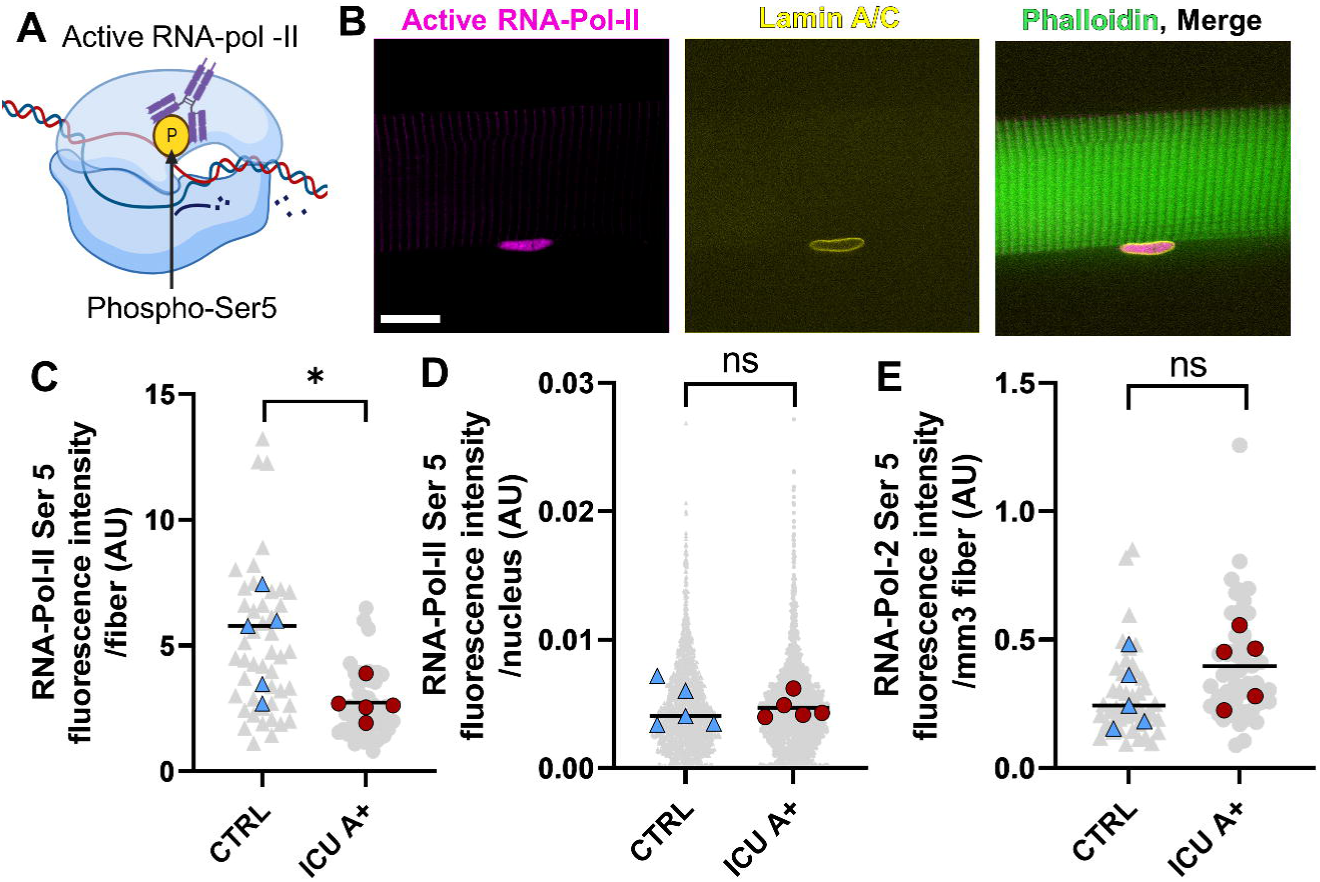
Total transcriptional activity is reduced in atrophic myofibers of ICU patients. **A**: Schematic of RNA-polymerase-II with phosphorylation at Serine 5 with antibody binding to phosphorylated Serine. This phosphorylation occurs when RNA-Pol-II starts transcribing DNA into mRNA. **B:** Representative images of RNA-Pol-II-Ser5 (magenta), lamin A/C (yellow), and phalloidin (green) labeling of a single Z-plane of a mounted single muscle fiber. Z-stacks were used to create 3D renders of fiber segments and total RNA-Pol-II Ser5 fluorescence intensity was measured within each myonucleus, using lamin A/C to segment all nuclei in 3D. Scale bar = 20 µm. **C:** Total RNA-Pol-II-Ser5 fluorescence intensity was calculated as the sum of the total RNA-Pol-II-Ser5 fluorescence intensity within each nucleus present within three Z-stacks generated from a single myofiber. CTRL *N = 5* patients, *n* = 48 myofibers; ICU A+ *N =* 5 patients, *n* = 48 myofibers. Each grey symbol represents the value of a single myofiber. Each colored symbol represents the mean value of a single patient. The black bar represents the mean value within the groups of patients. **D:** Total RNA-Pol-II-Ser5 fluorescence intensity per segmented nucleus. CTRL *N =* 5 patients, *n* = 2817 nuclei; ICU A+ *N = 5* patients, *n* = 2235 nuclei. Each grey symbol represents the value of a single nucleus. Each colored symbol represents the mean value of a single patient. The black bar represents the mean value within the groups of patients. **E.** Total RNA-Pol-II-Ser5 fluorescence intensity per myofiber volume was calculated as the sum of the total RNA-Pol-II-Ser5 fluorescence intensity within each nucleus present within the myofiber divided by the myofiber volume. CTRL *N = 5* patients, *n* = 48 myofibers; ICU A+ *N = 5* patients, *n* = 48 myofibers. Each grey symbol represents the value of a single myofiber. Each colored symbol represents the mean value of a single patient. The black bar represents the mean value within the groups of patients. Significance level was calculated using linear mixed models with the patients as the random factor. ICU A+ = ICU group with atrophy; CTRL = Control group; * denotes *p* < 0.05; ** denotes *p* < 0.01

### Diminished transcriptional activity of myofibers in the atrophic diaphragm of mechanically ventilated ICU patients

Myonuclear number has been shown to influence myonuclear transcription, with transcription levels increasing with decreasing nuclear number (29, 30). Thus, we aimed to determine whether changes in myonuclear number and morphology resulted in changes in myonuclear transcription in ICU patients. We hypothesized that diminished transcriptional activity contributes to atrophy in ICU patients. Furthermore, transcriptional activity may be downscaled with the reduced myonuclear domain size in ICU A+ patients, because the cellular volume to regulate is smaller. The transcriptional activity of each nucleus was determined by measuring the fluorescent intensity of phosphorylated Serine 5 on RNA-polymerase-2. This phosphorylation occurs shortly after the initiation of transcription, before the capping of the mRNA (31, 32) (Fig. 5A), and strongly correlates with transcriptional activity (33, 34). We measured the intensity of the immunofluorescence of activated RNA-polymerase-2 within each nucleus (Fig. 5B) from ten randomly selected myofibers of 5 patients from the ICU group and 5 controls (Table S7). Total transcriptional activity per fiber was calculated by summation of the fluorescence intensity of RNA-pol-II Ser5 labeling within each nucleus present in the myofiber and was on average almost two-fold lower in the ICU A+ group compared to the control group (Fig. 5C). This was explained by the lower myonuclear number in ICU myofibers, because the fluorescence intensity of RNA-Pol-II Ser5 per nucleus did not differ between the groups (Fig. 5D). After normalization to fiber volume, the difference in transcriptional activity per fiber was absent (Fig. 5E). Thus, based on these findings, insufficient transcriptional activity of individual nuclei is unlikely to contribute to atrophy, whereas the reduced myonuclear number in the ICU group with atrophy may reduce total transcriptional activity per myofiber.

### Decreased muscle stem cell number in the diaphragm of ICU patients

After muscles undergo atrophy that is accompanied by loss of myonuclei (Fig. 3) and transcriptional capacity (Fig. 5), myonuclear number could be restored by the contribution of satellite cells, the resident stem cells present adjacent to myofibers that contribute to myofiber homeostasis (35). Interestingly, muscle stem cell activity is particularly high in the diaphragm (36, 37). Thus, we aimed to investigate whether the muscle stem cell population in the atrophic diaphragm is affected, as this could further impair muscle recovery after atrophy. First, in our RNA-seq data set, we found a lower relative expression of *PAX7*, a transcript uniformly expressed in muscle stem cells (38), in both ICU groups compared to the control group (Fig. S1B). Next, immunofluorescent labeling of *PAX7* was performed to determine the number of muscle stem cells in diaphragm cross-sections (Fig. 6A). Patient characteristics of the groups used in these experiments are shown in Table S8. In ICU patients with atrophy, satellite cell content was reduced when normalized for either the number of myofibers present within the section (Fig. 6B) or when normalized for total section area (Fig. 6C). In conclusion, satellite cell content is reduced in the atrophic diaphragm of mechanically ventilated patients, potentially impeding the restoration of myonuclear number during recovery.

**Figure 6.**
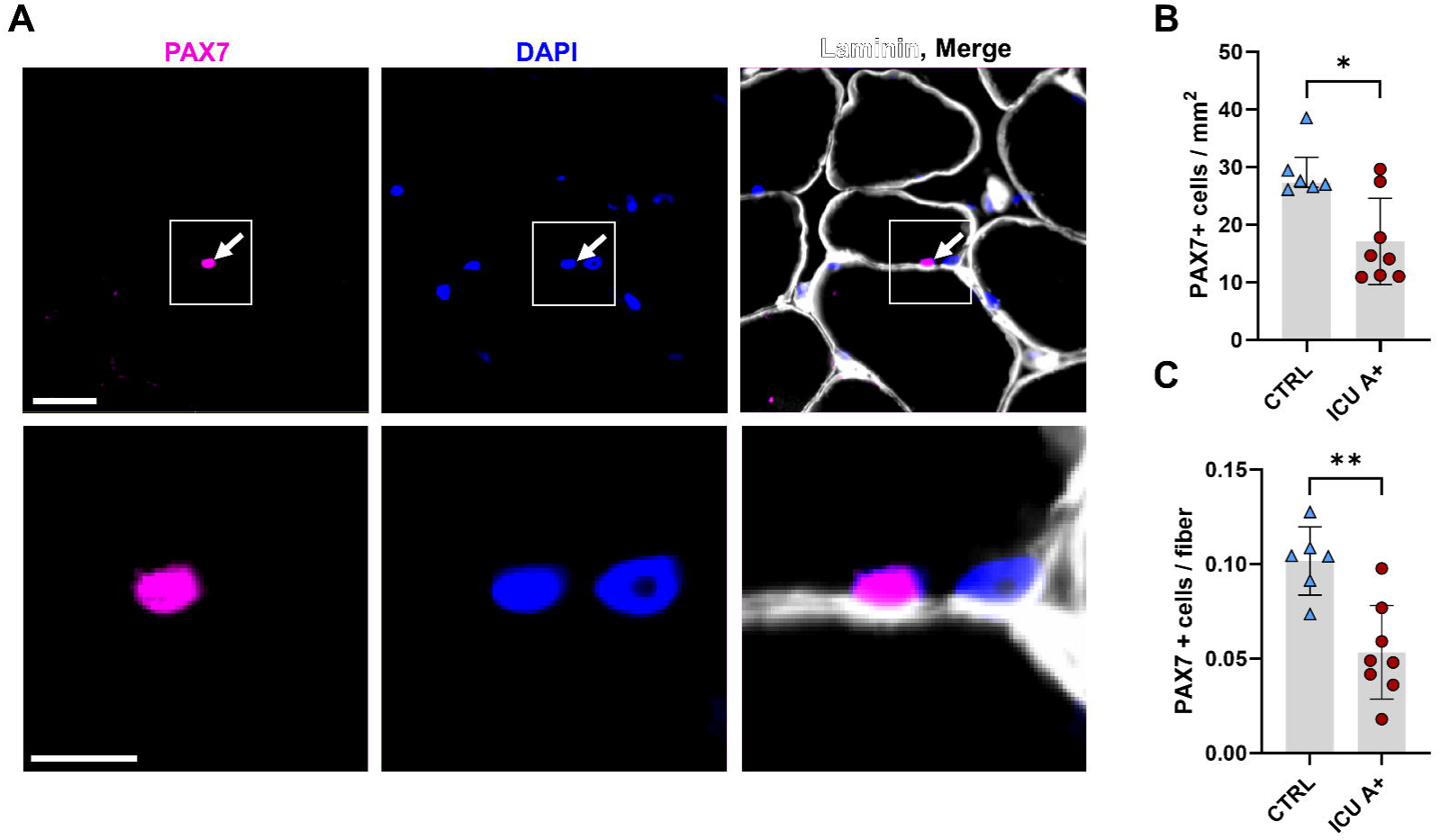
Decreased abundance of PAX7 positive cells in the atrophic diaphragm A: Representative images of diaphragm muscle cross-sections stained with PAX7 antibody, Laminin antibody, and DAPI. Nuclei with a PAX7-positive signal were designated as satellite cells. Scale bar = 50 µm in the top row and 10 µm in the bottom row. **B:** Number of PAX7 positive cells present in diaphragm muscle cross sections, normalized for section size. The grey bar represents the median [IQR] within the groups of patients. Each colored symbol represents a single patient. CTRL *N* = 6; ICU *N* = 8. Significance level was calculated using Mann-Whitney U test. **C:** Number of PAX7 positive cells present in diaphragm muscle cross sections, normalized for the number of myofibers. The grey bar represents the median [IQR] within the groups of patients, brackets represent interquartile ranges. Every colored symbol represents a single patient. CTRL *N* = 6; ICU *N* = 8. Significance level was calculated using a Mann-Whitney-U test. ICU = ICU group with atrophy, CTRL = Control group, * = *p*<0.05, ** = *p*<0.01

## Discussion

In this study, transcriptome profiling identified apoptotic pathway activation as a potential mechanism underlying critical illness-associated diaphragm weakness. Increased apoptosis of myonuclei was confirmed by using staining of cross-sections. Furthermore, this is the first study to reveal reduced myonuclear number in mechanically ventilated ICU patients. We show that myonuclear loss is associated with a reduction in transcriptional activity within myofibers. Having lost a significant number of myonuclei may put patients at a disadvantage during ventilator weaning and also may have long-term consequences after ICU discharge, because muscle growth and recovery is associated with myonuclear number (19, 20).

Reduced myonuclear number in atrophic myofibers implies that recovery of strength requires the addition of new myonuclei from satellite cells, instead of solely increasing protein synthesis to recover lost contractile material (Fig. 7) (39). Importantly, we discovered a reduced number of satellite cells in the diaphragm of ICU patients, possibly leading to further impairment of recovery after atrophy. Discovery of this underlying mechanism may pave the way for new treatments to prevent diaphragm atrophy.

**Figure 7.**
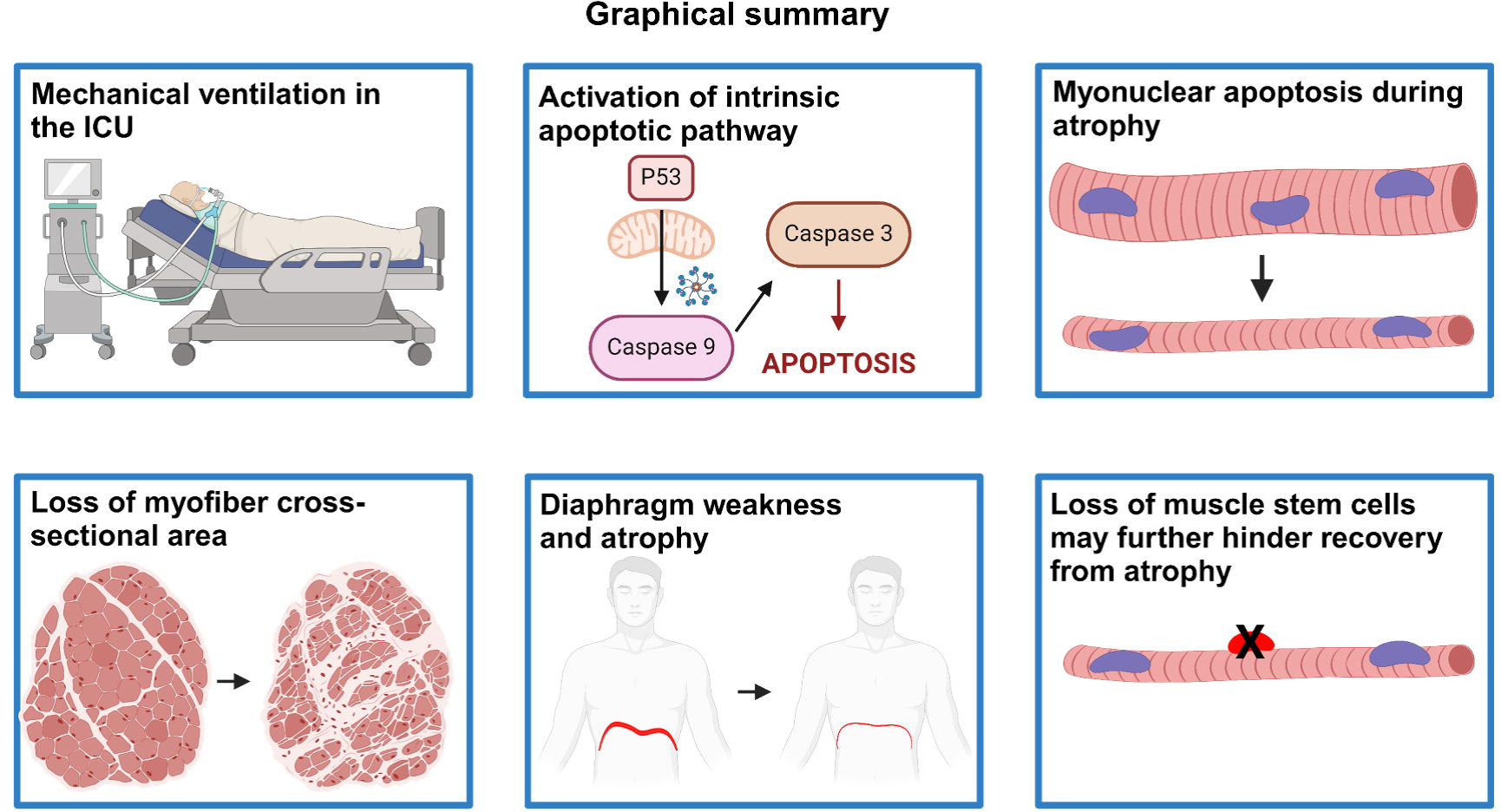
Graphical summary. Graphical summary of findings. Created using biorender.com.

### Myonuclear apoptosis in the diaphragm

The fate of myonuclei in muscles undergoing atrophy is subject to debate (40–42). The changes in myonuclear number and domain during atrophy likely depend on the type of atrophic stimulus (11, 43). Our observation that mechanical ventilation may induce caspase-3-mediated apoptosis of myonuclei is in accordance with the findings of a study investigating diaphragm atrophy in mechanically ventilated rats (6 and 12 h) (44). The authors reported no changes in myonuclear domain size, indicating atrophy that was proportional to myonuclear loss. In our study, the myonuclear domain was significantly smaller in the ICU group with established atrophy compared to controls, implying more or faster atrophy than myonuclear loss. There are several potential explanations for this discrepancy. First, we used 3D rendering of Z-stacks of confocal images to measure myonuclear domain volume within single myofibers, a more sensitive method compared to the myonuclear domain surface area (2D) measurements done on muscle cross-sections in the rodent study (15). Second, the patients in this study had a median ventilation duration of more than 60 hours, much longer than the 12 hours in the rodent study. In larger clinical studies, duration of ventilation was associated with the degree of diaphragm atrophy (45). The longer time on mechanical ventilation in our study may explain the reduction in myonuclear domain size, with atrophy exceeding myonuclear loss. Finally, unlike critically ill patients, the mechanically ventilated rodents were healthy. Common conditions in the ICU such as systemic inflammation and increased metabolic demand may independently cause muscle wasting, further accelerating atrophy (46). The pathophysiology of diaphragm atrophy has also been evaluated in mechanically ventilated brain-dead organ donors (7). Oxidative stress and mitochondrial dysfunction were suggested to activate the intrinsic pathway leading to caspase-3 activation and apoptosis (18). In diaphragm biopsies of mechanically ventilated ICU patients, a redox imbalance was proposed to underlie diaphragm atrophy in the absence of mitochondrial dysfunction or oxidative stress. This discrepancy may be due to the difference in clinical features between brain-dead organ donors and mechanically ventilated ICU patients.

In quadriceps muscle biopsies of ICU patients, we showed increased caspase-3 activity, albeit at a lower level than in the diaphragm, indicating caspase-3 activation before the onset of muscle atrophy and loss of myonuclei, independent of MV. Indeed, in the quadriceps of MV ICU patients, atrophy has been shown to occur later (7 days) after ICU admission (47).

In this study, we report a transcriptomic profile of the diaphragm of mechanically ventilated ICU patients. Main findings include the upregulation of 20 genes associated with the p53 pathway, as well as the upregulation of 16 genes associated with apoptosis. Because p53 is subject to redox regulation, a redox imbalance may activate the pathway (48). The p53 pathway was upregulated in various animal models of muscle atrophy. In mouse soleus muscle, after 48 of hind limb suspension, the p53 pathway was upregulated and the apoptotic index was increased (49). In a recent study in mechanically ventilated rabbits, the p53 pathway was upregulated and was hypothesized to contribute to diaphragm weakness via increased senescence (50). Interestingly, the p53 and apoptosis pathways were not upregulated in a transcriptomics study of the atrophic diaphragm of controlled mechanically ventilated rats (51). This demonstrates the disparity between animal models and clinical reality, further underlining the need to study pathophysiology in samples from patients.

Finally, 19 genes associated with the integrin pathway were upregulated in the diaphragm of ICU patients. This pathway is associated with the transmission of forces from the extracellular matrix to actin and has been shown to be upregulated after eccentric (lengthening) contractions (52). Eccentric contractions of the diaphragm may occur during ineffective efforts, premature cycling or reverse triggering, a ventilator asynchrony that has been shown to contribute to diaphragm dysfunction and myofiber injury in MV pigs with high respiratory effort (53, 54). Therefore, our data support a role for eccentric contractions in the pathophysiology of critical illness associated diaphragm weakness.

### Decreased number of satellite cells in the mechanically ventilated diaphragm

Our results show that the number of *PAX7* expressing cells is reduced in the diaphragm of mechanically ventilated ICU patients. This may be a direct effect of increased apoptosis, but we did not perform a separate experiment to quantify apoptotic satellite cells. Only a small fraction of cells in our cross-sections expressed *PAX7* (0.05-0.1%) Therefore, it is technically challenging to capture apoptotic satellite cells, especially because of diaphragm biopsy size limitations. Whether satellite cells are required for muscle regeneration depends on the type of atrophy, and on whether myonuclei are lost during atrophy (35, 55, 56). Nevertheless, lineage tracing studies demonstrated that satellite cell activity is particularly high in the diaphragm under non-diseased conditions (36, 37). In addition, there are interstitial stem cells that can contribute to myofibers that do not express *PAX7* (57–60). Future studies should determine the prevalence of non-satellite cell myogenic progenitors in the diaphragm of humans, and whether these cells are a significant source of myonuclei during diaphragm muscle homeostasis. Satellite cell content was reduced in the quadriceps of mechanically ventilated patients with sustained atrophy after 6 months, suggesting a crucial role for satellite cells during muscle regeneration in ICU patients (54). However, the exact role of satellite cells during recovery after diaphragm atrophy warrants further investigation.

Disturbed sarcomeric integrity is accepted as one of the mechanisms contributing to critical illness-associated diaphragm weakness (8). In non-diseased conditions, muscle can repair its sarcomeres, as damage occurs during both exercise and normal use. After sarcomeres are damaged, nuclear movement to a site of injury is required for repair (61, 62). During this process, myonuclei migrate toward damaged muscle sections to supply mRNA for sarcomeric proteins, allowing the reassembly of the contractile machinery (61). This mechanism may be disturbed in critical illness-associated diaphragm weakness because sarcomeric damage and reduced myonuclear number are present simultaneously.

### Clinical implications

The identification of intrinsic apoptotic pathway activation as a mechanism underlying diaphragm atrophy in MV ICU patients may open therapeutic venues to limit diaphragm weakness and weaning failure. The intrinsic apoptotic pathway plays a central role in many pathologies (63). Therefore, many inhibitors targeting the different components of this pathway have been developed (64). Especially caspase-3 inhibition has been investigated extensively, and was shown to prevent diaphragm atrophy in mechanically ventilated rodents (44, 65). To date, none have progressed beyond pre-clinical studies, probably due to functions of caspase-3 outside of the intrinsic apoptotic pathway (65). However, there are other strategies to limit apoptosis, such as limiting redox imbalance or inhibiting other constituents of the intrinsic apoptotic pathway, such as BAX or BAK (64, 66). Additionally, increasing the generation of new myonuclei by stimulating satellite cell activation may be a strategy to help diaphragm recovery after atrophy and speed up the weaning process.

Signaling pathway and immune modulators and various growth factors have been shown to augment satellite cell activation (67). More preclinical research is necessary to further explore these targets. Even though our data does not suggest that apoptotic pathway activation is limited to the diaphragm, we only observed myonuclear loss in the presence of myofiber atrophy, suggesting an important role for strategies to prevent atrophy such as diaphragm protective mechanical ventilation or diaphragm pacing (68, 69).

### Limitations

The population of ICU patients included in this is highly heterogeneous, with varying medical histories, reasons for admission, duration of mechanical ventilation and underlying pathophysiology. Therefore, it is not feasible to search for clinical predictors associated with myonuclear apoptosis or myofiber atrophy with our sample size. Nevertheless, this diverse group of patients does adequately reflect the general ICU population. Due to the invasiveness of taking diaphragm biopsies, the controls in this study are patients undergoing surgery for a pulmonary nodule. We cannot rule out an effect of the clinical status of the controls on the diaphragm, even though the in-and-exclusion criteria should minimize this. Finally, due to biopsy size limitations, not all experiments were performed on all biopsies.

## Conclusion

In mechanically ventilated patients in the ICU, myonuclear apoptosis is a pathophysiological mechanism underlying critical illness-associated diaphragm atrophy. Using a combination of advanced microscopy and molecular biology techniques, we identified p53 activation as the underlying pathway in ICU patients. Myonuclear loss in combination with a reduction of the satellite cell population may compromise recovery of the diaphragm, thereby contributing to weaning failure.

## Data Availability

All data produced in the present study are available upon reasonable request to the authors

