## Supplementary material for "Myonuclear apoptosis underlies diaphragm atrophy in mechanically ventilated ICU patients": Online supplement

*Patients, Diaphragm studies*

In this multicenter study, diaphragm muscle biopsies were taken from intensive care unit (ICU) patients receiving invasive mechanical ventilation (*ICU* patients, n=24) and patients undergoing elective lung surgery for early-stage lung malignancy without critical illness (*Control* patients, n=10). Exclusion criteria were chronic obstructive pulmonary disease (≥ GOLD stage III), congestive heart failure, neuromuscular diseases, chronic metabolic disorders, pulmonary hypertension, chronic use of corticosteroids (> 7.5mg/day for at least 3 months), and more than 10% weight loss within the last 6 months. The exclusion criteria were similar for all experimental groups. Patients' characteristics of the two groups are presented in Tables 1 and E1/E2. The biopsy protocol was approved by the institutional review board at Amsterdam UMC (location VUmc), the Netherlands. Patients were recruited in Amsterdam UMC and the Netherlands Cancer Institute-Antoni van Leeuwenhoek Hospital (both in Amsterdam, the Netherlands), and Medisch Spectrum Twente (Enschede, the Netherlands). Written informed consent was obtained from the patients or their legal representative. Biopsies were stored at -80ºC.

*Histology, myofiber cross-sectional area*

We used a previously described method for muscle specimen handling (1). Cryosections (8 µm thick, perpendicular to myofiber direction) were cut from the frozen biopsies and stained to study 1) myofiber type using myosin heavy chain antibodies; fast MHC (Table S10, #1), 2) myofiber cross-sectional area using wheat germ agglutinin (WGA, 1:25 diluted in PBS-0.5% BSA, Molecular Probes) staining. Myofiber cross-sectional area was obtained from whole cryosections.

*Single muscle fibers: myofiber volume and nuclear count*

Diaphragm biopsies were cut while submerged in liquid nitrogen. Segments of single diaphragm myofibers (length: 1-1.5 mm) were isolated in a relaxing solution at 5ºC (solution composition in supplementary Table 3. Single myofibers were attached to custom-made aluminum clips that were glued to glass coverslips ~2 mm apart, 6 pairs of aluminum clips per slide, and 2 slides per biopsy. A hydrophobic barrier was drawn using a PAP pen. The myofibers were fixed in 4% PFA for 15 minutes and permeabilized for 15 minutes and 1% triton X-100 in PBS. After blocking with 1% BSA in PBS, the fibers were incubated overnight with primary antibodies staining for Lamin A/C, MHC pan and fast MHC (Table S10, #2-4), at 4 C. The fibers were incubated overnight at 4C with secondary antibodies (Table S11, #1, #2, #3). Next, slides were incubated with DAPI (2μ/mL) for 5 minutes at room temperature and the samples were mounted in Vectashield (Vector Labs) and imaged the following week. The fibers were imaged using a CrestOptics X3 spinning disk confocal microscope using a Zeiss 63x magnification objective. Z-step size was 5 um, and three z-stacks with a thickness of the fiber thickness and a length of 208.5 μm were imaged. The Z-stacks were loaded into Imaris (Oxford Instruments) analysis software. Myofibers and nuclei were segmented using automatic thresholding using the MHC and Lamin A/C staining after a background subtraction pre-processing step. Each Z-stack was checked for wrongfully segmented nuclei and adjusted accordingly. Sarcomere length was determined in each z-stack using the MyofibrilJ plugin in FIJI (https://imagej.net/**MyofibrilJ**) on a single Z-slice of Myosin heavy chain staining. Nuclear counts were normalized for sarcomere length. Z-stacks of the Lamin A/C images were used to generate sum intensity projections using FIJI. The nuclei present in each fiber were segmented using the Stardist nuclear segmentation plugin for FIJI. Overlaying nuclei were excluded from the analysis. Wrinkling index of each nucleus was determined by calculating the standard deviation of the fluorescence intensity variation of Lamin A/C staining within the region of interest of each nucleus (2).

*Single muscle fibers: transcriptional activity*

Microscope slides with single myofibers attached to the coverslip were prepared as mentioned above, using the same blocking and permeabilization steps. The fibers were incubated overnight with primary antibodies (Phospho-RNA polymerase II CTD (Serine 5), fast MHC and Lamin A/C (Table S10, #1, #5, #6). Next, the fibers were incubated overnight at 4C with secondary antibodies (Table S11, #1, #2, #3). The next day, the fibers were incubated with pre-conjugated phalloidin (AF405) to stain actin. The fibers were mounted in Vectashield (Vector Labs) and imaged within the following week. The Z-stacks were loaded into Imaris analysis software, and the fibers and nuclei were segmented using the Lamin A/C and phalloidin staining after a background subtraction pre-processing step using automatic thresholding. Each Z-stack was checked for wrongfully segmented nuclei. Phospho-RNA polymerase II CTD (Serine 5) signal intensity was measured within each nucleus and used as a measure of transcriptional activity.

*Single muscle fibers: PCM-1 positive myonuclei*

Microscope slides with single myofibers attached to the coverslip were prepared as mentioned above, using the same blocking and permeabilization steps. The fibers were incubated overnight at 4C with primary antibody PCM-1 (Table S10, #7). Next, the fibers were incubated overnight at 4C with secondary antibody and AF647-conjugated phalloidin, 1:1000 Invitrogen. Next, slides were incubated with DAPI (2µg/mL) for 10 minutes at room temperature and the samples were mounted in Vectashield and imaged within the following week. Myofibers and myonuclei were segmented using Imaris software using automatic thresholding using phalloidin and PCM-1/DAPI staining after a background subtraction pre-processing step. The amount of PCM-1 positive nuclei in each fiber was determined automatically by the software and checked by a researcher blinded to the samples.

*TUNEL-assay*

Cryosections (8 µm thick, perpendicular to myofiber direction) were cut from the frozen biopsies. Apoptotic nuclei were identified by staining DNA breaks present in nuclei using a Click-iT™ plus TUNEL assay kits for in situ apoptosis detection, C10618, Invitrogen, according to the manufacturer's instructions. After TUNEL-staining, the sections were incubated overnight at 4°C with primary antibodies staining PCM-1 and Dystrophin (Table

S10, #7, #8). Next, the sections were incubated with secondary antibodies (Table S11, #5, #6) and incubated with DAPI, Invitrogen for 10 minutes. Per set of staining, a positive control was stained using DNAse-I to induce DNA breaks (supplemental figure 2). The sections were mounted with mowiol 4-88 (Calbiochem 475904) and imaged within the following week on a Zeiss AxioImager Fluorescent widefield microscope with a 20X objective. The images were analyzed using the FIJI plugin Stardist to segment nuclei. A blinded member of the study team manually counted TUNEL-positive nuclei. Each TUNEL-positive nucleus was then assessed for PCM-1 positive staining and positioning within the dystrophin barrier. Total myonuclear counts were determined using the FIJI plugin Stardist for automatic nuclear segmentation, then scored manually using PCM-1 staining. All automatically segmented images were visually inspected for accuracy and adjusted accordingly.

*Caspase-3 staining*

Cryosections (8 µm thick, perpendicular to myofiber direction) were cut from the frozen biopsies. The sections were incubated overnight at 4°C with primary antibodies staining PCM1, Cleaved caspase-3 and Laminin (Table S10 #9, #10, #11). Next, the sections were incubated with secondary antibodies (Table S11 #1, #3) and incubated with DAPI, Invitrogen for 10 minutes. The sections were mounted with mowiol and imaged within the following week on a Keyence BZ-X Fluorescent widefield microscope with a 20X objective. Sections were analyzed using FIJI with MuscleJ2 plugin (3). Activated caspase-3 positive nuclei were identified manually and assessed for PCM-1 positive staining and positioning within the laminin barrier. Analysis was performed by a researcher blinded to the samples.

*RNA-sequencing*

Diaphragm biopsies were cut while submerged in liquid nitrogen. A small piece was transferred to an Eppendorf tube and RNA was extracted using a kit (miRNAeasy kit, Qiagen) according to the manufacturer's instructions. Samples were sent to the Max-Planck-Institute for Heart and Lung Research in Bad Nauheim, Germany. RNA and library preparation integrity were confirmed with a LabChip Gx Touch 24 (Perkin Elmer). RNA integrity was assessed using a Agilent fragment analyzer, only samples with an RQN number >4.6 were further processed for sequencing. Libraries were prepared with 1µg of total RNA using SMARTer Stranded Total RNA Sample Prep Kit - HI Mammalian (Clontech). Sequencing was performed on a NextSeq500 (Illumina) with v2 chemistry, resulting in a minimum of 27M reads per library with a 75bp single-end setup. The resulting raw reads were assessed for quality, adapter content, and duplication rates with FastQC (http://www.bioinformatics.babraham.ac.uk/projects/fastqc). Trimmomatic version 0.39 was used to trim reads after a quality drop below a mean of Q20 in a window of 10 nucleotides. Only reads between 30 and 75 nucleotides were used in subsequent analyses. Trimmed and filtered reads were aligned against the human Ensembl genome version hg38 (GRC38.27) using STAR 2.6.1d with the parameter “-- outFilterMismatchNoverLmax 0.1” to increase the maximum ratio of mismatches to mapped length to 10%. The number of reads aligning to genes was counted using the featureCounts 1.6.5 tool from the Subread package. Only reads mapping at least partially inside exons were admitted and aggregated per gene. Reads overlapping multiple genes or aligning to multiple regions were excluded. Differentially expressed © 2020 American Medical Association. All rights reserved. genes were identified using DESeq2 version 1.26. The Ensemble annotation was enriched with UniProt data (release 06.06.2014) based on Ensembl gene identifiers (Activities at the Universal Protein Resource (UniProt)). GO-term analysis was performed with Metascape. The function of genes was referred to the “NCBI’s Gene” and “GeneCards” databases.

*PAX7 staining*

Cryosections (8 µm thick, perpendicular to myofiber direction) were cut from the frozen biopsies. Sections were air-dried and fixed with acetone. Next, the sections were incubated with PAX-7, laminin, and horse anti-mouse biotinylated (Table S10, #12, #13, Table S5, #7, #8), Then, sections were incubated with secondary antibodies (Table S11, #5, #8), and Hoechst (Life Technologies, H3569, 1:15000). The sections were mounted with mowiol and imaged within the following week on a Keyence BZ-X Fluorescent widefield microscope with a 20X objective. The PAX-7 positive cells were counted by hand.

*Statistical analysis*

Normality of the distribution of the studied variables was assessed visually on normal probability plots. Log transformation was performed if necessary. To compare the difference between ICU patients and control patients, student's t-test or Mann-Whitney-U was applied for non-repeat measurements; and linear mixed model with patients as the random factor was applied for measurements involving technical replicates in all human samples. For all linear mixed models, Greenhouse-Geisser correction was applied to adjust for potential lack of sphericity. For the comparison of three groups or more, one-way analysis of variance (ANOVA) or Kruskal-Wallis tests were performed with Tukey's or Dunn's post-hoc tests, depending on the distribution of the data. We used a two-sided significance level of 5% for all analyses. Unless otherwise noted, data are expressed as mean (± standard error), median [interquartile range], or frequencies (percentage), as appropriate.

**Supplemental figure legends**

**Figure S1. Relative expression levels of genes associated with the P53 pathway**

**A:** Relative expression levels of genes associated with the P53 pathway determined with RNA-sequencing. *p* -values were adjusted for repeated testing. CTRL *N* = 8; ICU *N* = 17. **B:** Relative expression levels of PAX7. CTRL *N* = 8; ICU *N* = 17. The ICU groups with and without atrophy were pooled to calculate the significance level. *p* -values were adjusted for repeated testing. CTRL = Control group, ICU = Intensive care patients

**Figure S2. Representative images of DNAse-treated muscle cross-section after TUNEL staining**

Note that every nucleus has a positive TUNEL signal after inducing DNA breaks with DNAse treatment. Scale bar is 50 µm.

**Figure S3. Apoptotic index of non-myonuclei**

**A:** Quantification of TUNEL index for non-myonuclei. The grey bar represents the median value within the groups of patients, brackets represent interquartile ranges. Each colored symbol represents the value of a single patient. ICU A+ *N* = 14; ICU A− *N* = 7; CTRL; *N* = 11. Significance level was calculated using Kruskal-Wallis test. **B:** Quantification of activated caspase-3 index for non-myonuclei. The grey bar represents the median value within the groups of patients. Each colored symbol represents the value of a single patient. ICU A+ *N* =

7; ICU A− $N$ = 6; CTRL; $N$ = 7 Significance level calculated T-test or Mann-Whitney U test, depending on distribution of the data.

**Figure S4. Increased Caspase-3 index in Quadriceps biopsies of MV patients. A:** Representative images of vastus lateralis muscle cross-sections stained with Cleaved caspase-3 antibody, PCM1 antibody, Laminin antibody and DAPI. Nuclei with a cleaved-Caspase-3 positive signal were designated as apoptotic myonuclei when they were PCM1 positive and were located within the laminin barrier. Top scale bar is 50μm, bottom scale bar is 20 μm. **B:** Quantification of activated caspase-3 index**,** calculated as the percentage of activated caspase-3-positive myonuclei. Total myonuclear count was determined by counting PCM1-positive nuclei. The grey bar represents the mean value within the groups of patients, brackets represent the standard deviation. Each colored symbol represents the value of a single patient. ICU $N$ = 10; CTRL $N$ = 5. The significance level was calculated using an unpaired T-test. **C:** Quantification of activated caspase-3 index for non-myonuclei. The grey bar represents the mean value within the groups of patients, brackets represent the standard deviation. Each colored symbol represents the value of a single patient. **D:** Mean myofiber CSA The grey bars represent the mean value of the groups. Colored symbols represent the value per patient. ICU $N$ = 10; CTRL $N$ = 5. The significance level was calculated using unpaired T-test. ICU; N = 10, CTRL; N = 5. The significance level was calculated using unpaired t-test. ICU = ICU group, CTRL = Control group, * = $p$<0.05, ** = $p$<0.01

**Figure S5. Comparison of activated-caspase-3 index between quadriceps and diaphragm biopsies.**

**A:** Quantification of activated caspase-3 index in all ICU patients**,** calculated as the percentage of activated caspase-3-positive myonuclei. Total myonuclear count was determined by counting PCM1-positive nuclei. The grey bar represents the median value within the groups of patients. Each colored symbol represents the value of a single patient. ICU Quad *N* = *10*; ICU A− Dia *N* = 6; ICU A+ *N* = 7. Significance level was calculated using Kruskal-Wallis test. **B:** Quantification of activated caspase-3 index in all control patients**,** calculated as the percentage of activated caspase-3-positive myonuclei. Total myonuclear count was determined by counting PCM1-positive nuclei. The grey bar represents the median value within the groups of patients. Each colored symbol represents the value of a single patient. CTRL Quad *N* = 5; CTRL Dia *N* = 7. Significance level calculated with unpaired t-test.

**Figure S6. Proportion of PCM-1 + nuclei within manually isolated single myofibers**

**A:** Representative images of a single myofiber stained with DAPI (blue) and PCM1 (yellow). Scale bar is 20 µm. Arrow indicates PCM1-negative nucleus. Note the PCM1 negative nucleus indicated with the white arrow. **B:** Proportion of PCM1 + nuclei within manually isolated myofibers from controls *N* = 3. **C:** Proportion of PCM1 + nuclei within manually isolated myofibers from ICU patients *N* = 3.

**Figure S7. Myofiber volume, myonuclear number and myonuclear domain of isolated myofibers in two ICU groups**

**A:** Quantification of myofiber volume, calculated as volume per mm fiber was normalized to a sarcomere length of 2.5 µm. Every grey dot represents the value of a single muscle fiber, and the colored symbols represent the mean values of a single patient. Slow-twitch fibers: ICU *N* = 24, *n* = 134; CTRL *N* = 10, *n* = 48. Fast-twitch fibers: ICU *N* = 24, *n* = 104; CTRL *N* = 10, *n* = 46. **C:** Quantification of myonuclear number. Every grey dot represents the value of a single muscle fiber and the colored symbols represent the mean values of a single patient. Slow-twitch fibers: ICU *N* = 24, *n* = 134; CTRL *N* = 10, *n* = 48. Fast-twitch fibers: ICU *N* = 24, *n* = 104; CTRL *N* = 10, *n* = 46. **D:** Quantification of myonuclear domain size. Every grey dot represents the value of a single muscle fiber and the colored symbols represent the mean values of a single patient. Significance levels were calculated using linear mixed models with the patients as the random factor. Black bars indicate the median of the whole group. Slow-twitch fibers: ICU $N = 24$, $n = 134$; CTRL $N = 10$, $n = 48$. Fast-twitch fibers: ICU $N = 24$, $n = 104$; CTRL $N = 10$, $n = 46$. ICU = ICU group; CTRL = Control group. * denotes $p < 0.05$; ** denotes $p < 0.01$. N = number of patients, n = number of analyzed myofibers.

**Figure S8. Myonuclear content of diaphragm and quadriceps cross-sections**

**A:** Representative images of diaphragm cross-sections with DAPI (blue) PCM1 (yellow) and Dystrophin (grey). Scale bar is 50 µm. **B:** Myonuclear number per myofiber. The grey bars represent the mean value of the groups. Colored symbols represent the value per patient. CTRL = 8; ICA A- = 7; ICA A+ = 11. Statistics are performed using one-way ANOVA. Post-hoc testing with Tukey's test. CTRL = Control group, ICU A+ = Intensive care patients with myofiber atrophy, ICU A− = Intensive care patients without atrophy, **C:** Myonuclear number per myofiber. The grey bars represent the mean value of the groups. Colored symbols represent the value per patient. CTRL QUAD = control group quadriceps biopsies, ICU QUAD = ICU group, quadriceps biopsies. Statistics are performed using a T-test. ** = $p<0.01$, ****= $p<0.0001$

**Figure S9. Scatterplots of nuclear number vs. myofiber cross-sectional area and myonuclear domain vs. cross-sectional area**

**A:** Number of nuclei plotted against fiber CSA of every fiber of the ICU A+ (dark red) and ICU A− (light red) groups, compared with the CTRL (blue) group. Slow-twitch fibers: ICU A+ $N = 14$, $n = 84$; ICU A− $N = 10$, $n = 50$; CTRL $N = 10$, $n = 48$. Fast-twitch fibers: ICU A+ $N = 14$, $n = 56$; ICU A− $N = 10$, $n = 48$; CTRL $N = 10$, $n = 46$. R-squared and $p$-value were calculated using simple linear regression. The slope and elevation were compared between the groups using Analysis of Covariance (ANCOVA). **B:** Myonuclear domain plotted against fiber CSA of every fiber of the ICU A+ (dark red) and ICU A− (light red) groups, compared with the CTRL (blue) group. R-squared and $p$-value were calculated using simple linear regression. The slope and elevation were compared between the groups using Analysis of Covariance (ANCOVA). Slow-twitch fibers: ICU A+ *N* = 14, *n* = 84; ICU A− *N* = 10, *n* = 50; CTRL *N* = 10, *n* = 48. Fast-twitch fibers: ICU A+ *N* = 14, *n* = 56; ICU A− *N* = 10, *n* = 48; CTRL *N* = 10, *n* = 46.

ICU A+ = ICU group with atrophy, ICU A− = ICU group without atrophy, CTRL = Control group, MND = Myonuclear domain, * = $p<0.05$, ** = $p<0.01$

**Figure S10. Myonuclear ellipticity and sphericity**

**A:** Ellipticity of nuclei within single myofibers. Every grey dot represents the value of a single nucleus. Every colored symbol represents the mean value per patient. The black bars represent the mean value per group of patients. The significance level was determined using linear mixed models with the patients as the random factor. CTRL *N* = 10 patients, *n* = 6270 nuclei; ICU A+ *N* = 14 patients; *n* = 5463 nuclei; ICU A− *N* = *10* patients; *n* = 6330 nuclei. **B:** sphericity of nuclei within single myofibers. Every grey dot represents the value of a single nucleus. Every colored symbol represents the mean value per patient. The black bars represent the mean value per group of patients. The significance level was determined using linear mixed models with the patients as the random factor. CTRL *N* = 10 patients, *n* = 6270 nuclei; ICU A+ *N* = 14 patients; *n* = 5463 nuclei; ICU A− *N* = *10* patients; *n* = 6330 nuclei.

ICU A+ = ICU group with atrophy, ICU A− = ICU group without atrophy, CTRL = Control group

### Supplementary tables.

**Supplementary table 1. Experiments performed per diaphragm biopsy.**

| Subject # | Group | Nuclear number | Transcr. activity | RNA-seq | Apoptosis TUNEL | Apoptosis Casp3 | PAX 7 |
|---|---|---|---|---|---|---|---|

| | | and<br><br>MND | | | | | |
|---|---|---|---|---|---|---|---|
| **25** | ICU A+ | 1 | 0 | 1 | 0 | 0 | 0 |
| **46** | ICU A+ | 1 | 0 | 0 | 0 | 0 | 0 |
| **42** | ICU A+ | 0 | 0 | 1 | 0 | 0 | 0 |
| **50** | ICU A+ | 0 | 0 | 0 | 1 | 0 | 0 |
| **52** | ICU A+ | 1 | 0 | 1 | 0 | 0 | 0 |
| **64** | ICU A+ | 0 | 0 | 1 | 1 | 0 | 1 |
| **72** | ICU A+ | 1 | 0 | 0 | 1 | 1 | 1 |
| **73** | ICU A+ | 1 | 1 | 0 | 1 | 0 | 1 |
| **74** | ICU A+ | 1 | 1 | 1 | 1 | 1 | 1 |
| **78** | ICU A+ | 1 | 0 | 0 | 1 | 1 | 0 |
| **80** | ICU A+ | 1 | 0 | 0 | 1 | 1 | 0 |
| **82** | ICU A+ | 1 | 0 | 1 | 1 | 1 | 0 |
| **89** | ICU A+ | 1 | 1 | 0 | 0 | 0 | 1 |
| **91** | ICU A+ | 1 | 0 | 0 | 1 | 1 | 0 |
| **94** | ICU A+ | 1 | 0 | 1 | 1 | 0 | 0 |
| **99** | ICU A+ | 1 | 1 | 1 | 1 | 1 | 1 |
| **100** | ICU A+ | 1 | 1 | 0 | 1 | 1 | 1 |
| **101** | ICU A+ | 0 | 0 | 1 | 0 | 0 | 0 |
| **3** | ICU A - | 0 | 0 | 0 | 1 | 0 | 0 |
| **12** | ICU A - | 1 | 0 | 0 | 1 | 1 | 0 |
| **15** | ICU A - | 1 | 0 | 0 | 0 | 0 | 0 |
| **22** | ICU A - | 1 | 0 | 0 | 1 | 1 | 0 |
| **27** | ICU A - | 1 | 0 | 1 | 0 | 0 | 0 |
| **38** | ICU A - | 0 | 0 | 1 | 0 | 0 | 0 |
| **57** | ICU A - | 0 | 0 | 1 | 0 | 0 | 0 |

| | | | | | | | |
|---|---|---|---|---|---|---|---|
| **66** | ICU A - | 1 | 0 | 0 | 1 | 1 | 0 |
| **70** | ICU A - | 1 | 0 | 1 | 1 | 1 | 0 |
| **77** | ICU A - | 1 | 0 | 1 | 1 | 1 | 0 |
| **83** | ICU A - | 1 | 0 | 0 | 1 | 0 | 0 |
| **95** | ICU A - | 0 | 0 | 1 | 0 | 0 | 0 |
| **96** | ICU A - | 1 | 0 | 0 | 1 | 1 | 0 |
| **97** | ICU A - | 0 | 0 | 1 | 0 | 0 | 0 |
| **6** | CTRL | 0 | 0 | 1 | 0 | 0 | 0 |
| **7** | CTRL | 0 | 0 | 1 | 0 | 0 | 0 |
| **8** | CTRL | 1 | 0 | 0 | 1 | 1 | 0 |
| **41** | CTRL | 1 | 1 | 1 | 0 | 1 | 1 |
| **43** | CTRL | 1 | 0 | 1 | 1 | 1 | 1 |
| **45** | CTRL | 0 | 0 | 1 | 0 | 0 | 0 |
| **48** | CTRL | 1 | 1 | 0 | 1 | 1 | 1 |
| **49** | CTRL | 1 | 1 | 0 | 1 | 1 | 1 |
| **55** | CTRL | 1 | 0 | 0 | 1 | 1 | 1 |
| **56** | CTRL | 1 | 0 | 0 | 1 | 0 | 1 |
| **58** | CTRL | 1 | 1 | 1 | 1 | 1 | 1 |
| **60** | CTRL | 0 | 0 | 1 | 0 | 0 | 0 |
| **62** | CTRL | 1 | 1 | 1 | 1 | 0 | 0 |
| **63** | CTRL | 1 | 0 | 0 | 0 | 0 | 1 |
| **79** | CTRL | 0 | 0 | 1 | 0 | 0 | 0 |

ICU A+ = ICU group with atrophy, ICU A− = ICU group without atrophy, CTRL = Control group.

**Supplementary table 2. Patient characteristics, RNA-sequencing experiment**

| | Control (n=9) | ICU (n=17) | P-value |
|---|---|---|---|
| **Age (years)** | 64 [51-71] | 66 [42-75] | 0.968 |
| **M (%)** | 7 (78) | 8 (47) | 0.132 |
| **BMI ($Kg/m^2$)** | 24 [23-30] | 25 [23-28] | 0.804 |
| **APACHE-3** | - | | - |
| **Ventilation (hours)** | 1.3 [0.9-2] | 58 [42-169] | <0.001 |
| **Myofiber CSA ($\mu m^2$)** | 3021 [2476-3542] | 2067 [1729-3039] | 0.099 |

*BMI = Body Mass Index APACHE = Acute Physiology And Chronic Health Evaluation, CSA = Cross-Sectional Area. Data shown as Median [IQR]. P-values of continuous data calculated with unpaired T-test or Mann-Whitney-U test, depending on distribution of the data. P-values of categorical data calculated with Chi-squared test.*

**Supplementary table 3. Patient characteristics, TUNEL experiment**

| | Control (n=8) | ICU (n=18) | P-value |
|---|---|---|---|
| **Age (years)** | 67 [58-73] | 66 [50-72] | 0.676 |
| **M (%)** | 3 (38) | 5 (28) | 0.620 |
| **BMI ($Kg/m^2$)** | 27 [24-29] | 27 [23-30] | 0.380 |
| **APACHE-3** | - | 66 [26-104] | - |
| **Ventilation (hours)** | 1.5 [0.9-2.0] | 99 [56-205] | <0.001 |
| **Myofiber CSA($\mu m^2$)** | 2147 [1624-2816] | 1563 [1064-3366] | 0.812 |

*BMI = Body Mass Index APACHE = Acute Physiology And Chronic Health Evaluation, CSA = Cross-Sectional Area. Data shown as Median [IQR]. P-values of continuous data calculated with unpaired T-test or Mann-Whitney-U test, depending on distribution of the data. P-values of categorical data calculated with Chi-squared test.*

**Supplementary table 4. Patient characteristics, Caspase-3 staining experiment.**

| | Control (n=7) | ICU (n=14) | P-value |
|---|---|---|---|
| **Age (years)** | 66 [60-71] | 62 [47-70] | 0.456 |
| **M (%)** | 3 (57) | 5 (38) | 0.751 |
| **BMI (Kg/m$^2$)** | 26 [24-28] | 25 [22-28] | 0.864 |
| **APACHE-3** | - | 66 [37-96] | - |
| **Ventilation (hours)** | 1.0 [0.9-1.8] | 65 [36-205] | <0.001 |
| **Myofiber CSA (μm$^2$)** | 2501 [1720-3024] | 1684 [1064-3464] | 0.502 |

*BMI = Body Mass Index APACHE = Acute Physiology And Chronic Health Evaluation, CSA = Cross-Sectional Area. Data shown as Median [IQR]. P-values of continuous data calculated with unpaired T-test or Mann-Whitney-U test, depending on distribution of the data. P-values of categorical data calculated with Chi-squared test.*

**Supplementary table 5. Patient characteristics, quadriceps experiment**

| | Control (n=5) | ICU quadriceps (n=10) | P-value |
|---|---|---|---|
| **Age (years)** | 51 [21-58] | 63 [32-82] | 0.148 |
| **M (%)** | 4 (80) | 9 (90) | 0.591 |
| **BMI (Kg/m$^2$)** | 25 [23-28] | 23 [22-29] | 0.864 |
| **APACHE-4** | - | 97 [73-130] | - |
| **Ventilation (hours)** | - | 39 [17-50] | - |
| **Myofiber CSA (μm$^2$)** | 6509 [4828-9339] | 4438 [3723-5816] | 0.199 |

*BMI = Body Mass Index APACHE = Acute Physiology And Chronic Health Evaluation, CSA = Cross-Sectional Area. Data shown as Mean ± SD or Median [IQR], depending on distribution of the data. P-values of continuous data calculated with unpaired T-test or Mann-Whitney-U test, depending on distribution of the data. P-values of categorical data calculated with Chi-squared test.*

**Supplementary table 6. PCM-1 staining of manually isolated myofibers.**

| | ICU (n=3) | Control (n=3) | P-value |
|---|---|---|---|
| **Age** | 71 [61-75] | 64 [61-68] | 0.376 |
| **Male (%)** | 2 (60) | 2 (60) | 1.0 |
| **BMI** | 25 [20-28] | 31 [29-33] | 0.100 |
| **APACHE** | 87 [57-149] | - | - |
| **Ventilation (hours)** | 45 [24-84] | 1.0 [0.8-2.0] | 0.400 |
| **Myofiber CSA ($\mu m^2$)** | 1065 [530-1628] | 3128 [2437-3132] | 0.100 |

*BMI = Body Mass Index APACHE = Acute Physiology And Chronic Health Evaluation, CSA = Cross-Sectional Area*

**Supplementary table 7. Patient characteristics, transcriptional activity of nuclei experiment**

| | Control (n=5) | ICU (n=5) | P-value |
|---|---|---|---|
| **Age** | 64 [61-68] | 62 [61-71] | 1.0 |
| **Male (%)** | 3 (60) | 3 (60) | 1.0 |
| **BMI** | 29 [27-31] | 25 [20-25] | 0.032 |
| **APACHE-3** | - | 57 [53-87] | - |
| **Ventilation (hours)** | 1.0 [0.6-1.5] | 63 [45-84] | 0.008 |
| **Myofiber CSA ($\mu m^2$)** | 2501 [1975-3129] | 1318 [797-17] | 0.032 |

*BMI = Body Mass Index APACHE = Acute Physiology And Chronic Health Evaluation, CSA = Cross-Sectional Area. Data shown as Median [IQR]. P-values of continuous data calculated with unpaired T-test or Mann-Whitney-U test, depending on distribution of the data. P-values of categorical data calculated with Chi-squared test.*

**Supplementary table 8. Patient characteristics, PAX-7 staining experiment.**

| | Control (n=8) | ICU (n=8) | P-value |
|---|---|---|---|
| **Age (years)** | 64 [58-73] | 69 [60-73] | 0.798 |
| **M (%)** | 5 (63) | 5 (63) | 1.0 |
| **BMI (Kg/m$^2$)** | 28 [25-30] | 25 [22-29] | 0.328 |
| **APACHE-3** | - | 72 [15-124] | - |
| **Ventilation (hours)** | 1.3 [0.8-1.5] | 74 [49-131] | <0.001 |
| **Myofiber CSA (µm$^2$)** | 2180 [1667-2856] | 1192 [952-1709] | 0.015 |

*BMI = Body Mass Index  APACHE = Acute Physiology And Chronic Health Evaluation, CSA = Cross-Sectional Area. Data shown as Median [IQR]. P-values of continuous data calculated with unpaired T-test or Mann-Whitney-U test, depending on distribution of the data. P-value of categorical data calculated with Chi-squared test.*

**Supplementary table 9. Composition of solutions**

| Solutions | Composition |
|---|---|
| **Relaxing solution** | 100 mM BES, 14.5 mM creatine phosphate, 6.97 mM EGTA, 40.76 mM K-propionate, 6.48 mM $MgCl_2$, 5.89 mM $Na_2$-ATP, and low concentration of freshly added protease inhibitors. |

**Supplementary table 10. Primary antibodies**

| # | Antigen | Name | Type | Dilution | Source | Code |
|---|---|---|---|---|---|---|
| 1 | MYH 1 | MY-32 | Mouse mono IgG1 | 1:50 | Abcam | ab51263 |
| 2 | Lamin A/C | E1 | Mouse mono IgG1 | 1:200 | Santa Cruz | SC-376248 |
| 3 | MYH pan | A4.1025 | Mouse mono IgG2a | 1:600 | DSHB | A4.1025 |
| 4 | MYH 1 | MY-32 | Rabbit poly IgG | 1:1000 | Abcam | ab91506 |
| 5 | RNA-pol-II (Ser5) | 4H8 | Mouse mono IgG1 | 1:100 | Thermo | MA1-46093 |
| 6 | Lamin A/C | 4C11 | Mouse mono IgG2a | 1:200 | Cell signaling | 4777S |
| 7 | PCM1 | HPA023370 | Rabbit poly IgG | 1:2500 | Sigma | HPA023370 |
| 8 | Dystrophin | MANDRA1 7A10 | Mouse mono IgG1 | 1:400 | DSHB | MANDRA1 7A10 |
| 9 | PCM1 | G6 | Mouse mono IgG1 | 1:200 | Santa Cruz | sc398365 |
| 10 | Cleaved Casp-3 | Asp-175 | Rabbit poly IgG | 1:400 | Cell signaling | 9661 |
| 11 | Laminin | D18 | Mouse mono IgG2a | 1:200 | DSHB | D18 |
| 12 | PAX7 | PAX7 | Mouse mono IgG1 | 1:15 | DSHB | PAX7 |
| 13 | Laminin | L9393 | Rabbit poly IgG | 1:500 | Sigma | L9393) |

**Supplementary table 11. Secondary antibodies**

| # | Reactivity | Animal | Type | Dilution | Source | Code |
|---|---|---|---|---|---|---|
| 1 | Mouse IgG1 | Goat | AF 555 | 1:100 | Invitrogen | A21127 |
| 2 | Mouse IG2a | Goat | AF 488 | 1:100 | Invitrogen | A21131 |
| 3 | Rabbit IgG (H+L) | Donkey | AF647 | 1:100 | Invitrogen | A21447 |

| | | | | | | |
|---|---|---|---|---|---|---|
| 4 | Rabbit IgG (H+L) | Donkey | AF555 | 1:100 | Invitrogen | A21432 |
| 5 | Rabbit IgG (H+L) | Donkey | AF488 | 1:100 | Invitrogen | A11055 |
| 6 | Mouse IgG (H+L) | Donkey | AF647 | 1:100 | Invitrogen | A31574 |
| 7 | Mouse IgG | Horse | Biotinylated | 1:50 | Vectorlabs | BA2000 |
| 8 | Streptavidin | - | AF594 | 1:500 | Thermo | S32356 |

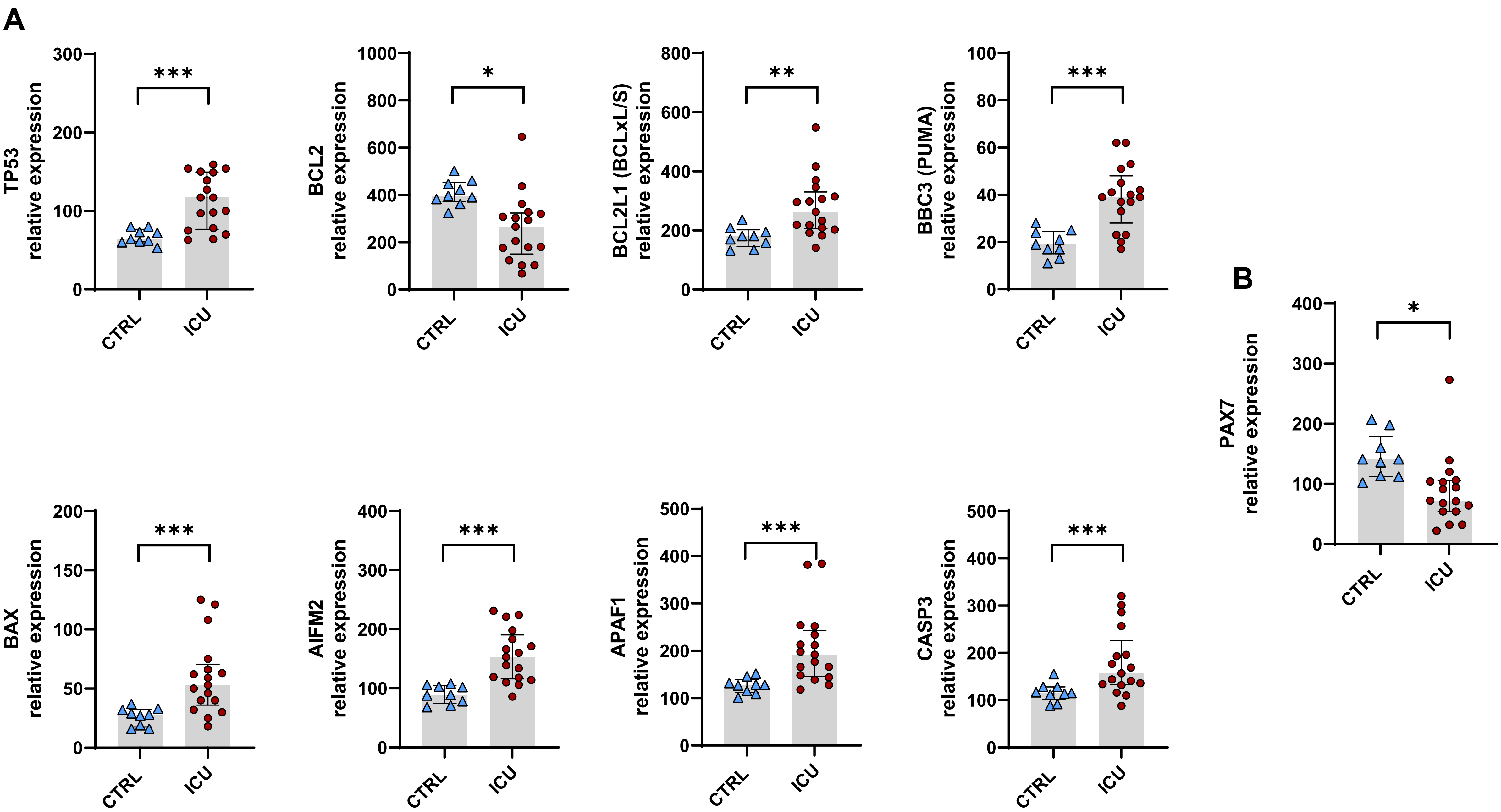
A
TP53 relative expression
BCL2 relative expression
BCL2L1 (BCLxL/S) relative expression
BBC3 (PUMA) relative expression
BAX relative expression
AIFM2 relative expression
APAF1 relative expression
CASP3 relative expression
B
PAX7 relative expression
CTRL
ICU
***
*
**

Figure S1

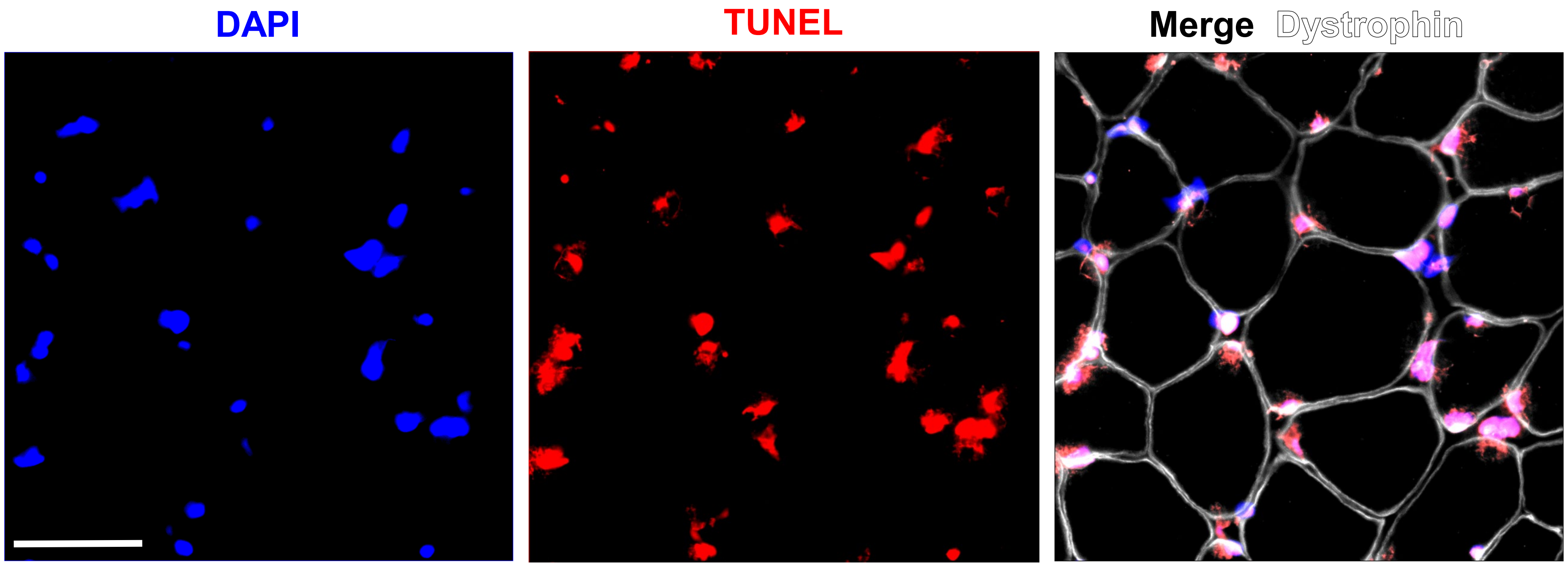


Figure S2

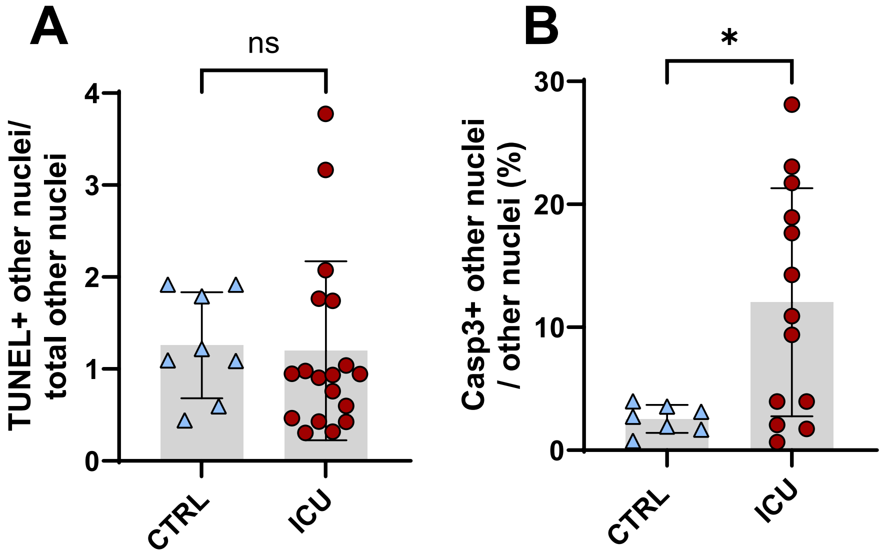
A
ns
TUNEL+ other nuclei/
total other nuclei
4
3
2
1
0
CTRL
ICU
B
*
Casp3+ other nuclei
/ other nuclei (%)
30
20
10
0
CTRL
ICU

**Figure S3**

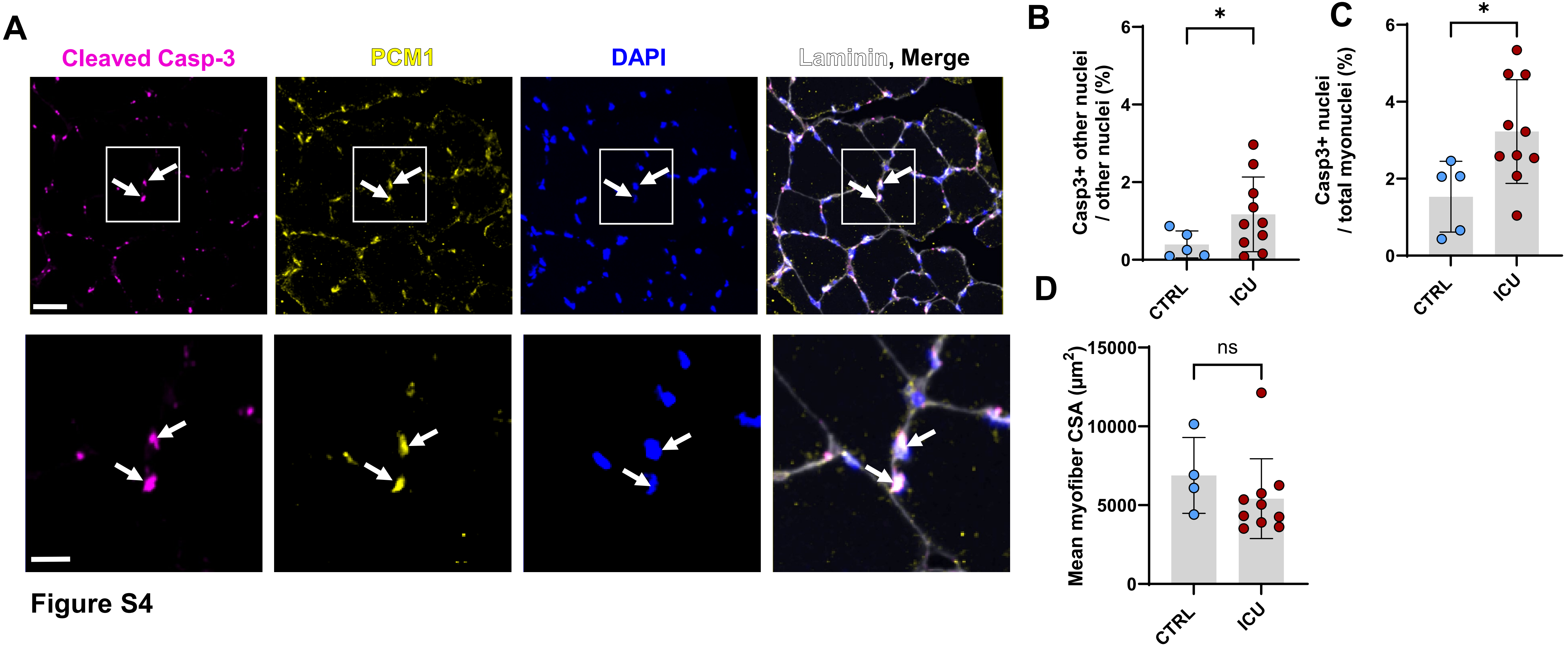
A
Cleaved Casp-3
PCM1
DAPI
Laminin, Merge
B
Casp3+ other nuclei / other nuclei (%)
*
CTRL
ICU
C
Casp3+ nuclei / total myonuclei (%)
*
CTRL
ICU
D
Mean myofiber CSA (μm2)
ns
CTRL
ICU

Figure S4

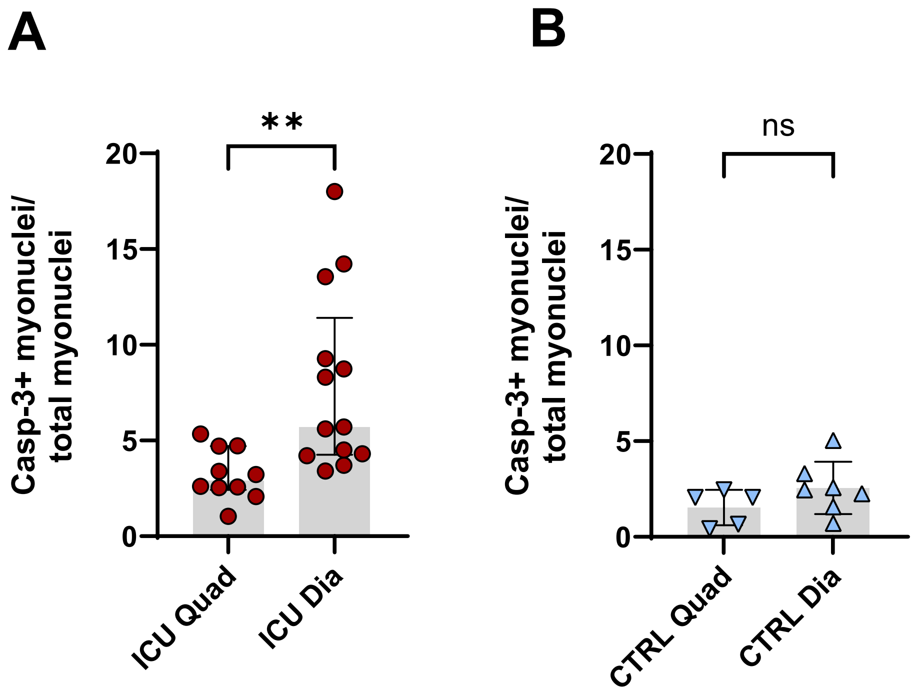
A
B
**
ns
Casp-3+ myonuclei/
total myonuclei
20
15
10
5
0
ICU Quad
ICU Dia
CTRL Quad
CTRL Dia

**Figure S5**

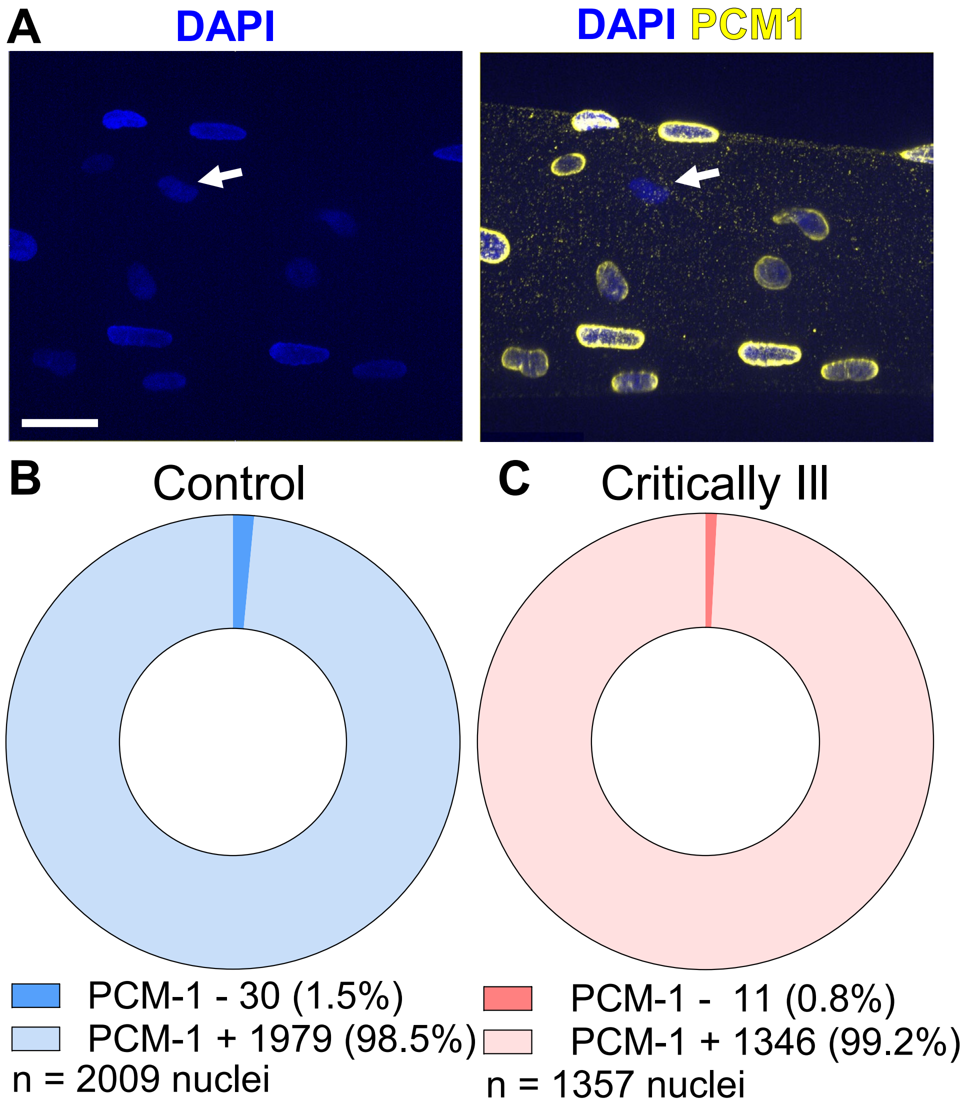
A
DAPI
DAPI PCM1
B
Control
C
Critically Ill
PCM-1 - 30 (1.5%)
PCM-1 + 1979 (98.5%)
n = 2009 nuclei
PCM-1 - 11 (0.8%)
PCM-1 + 1346 (99.2%)
n = 1357 nuclei

**Figure S6**

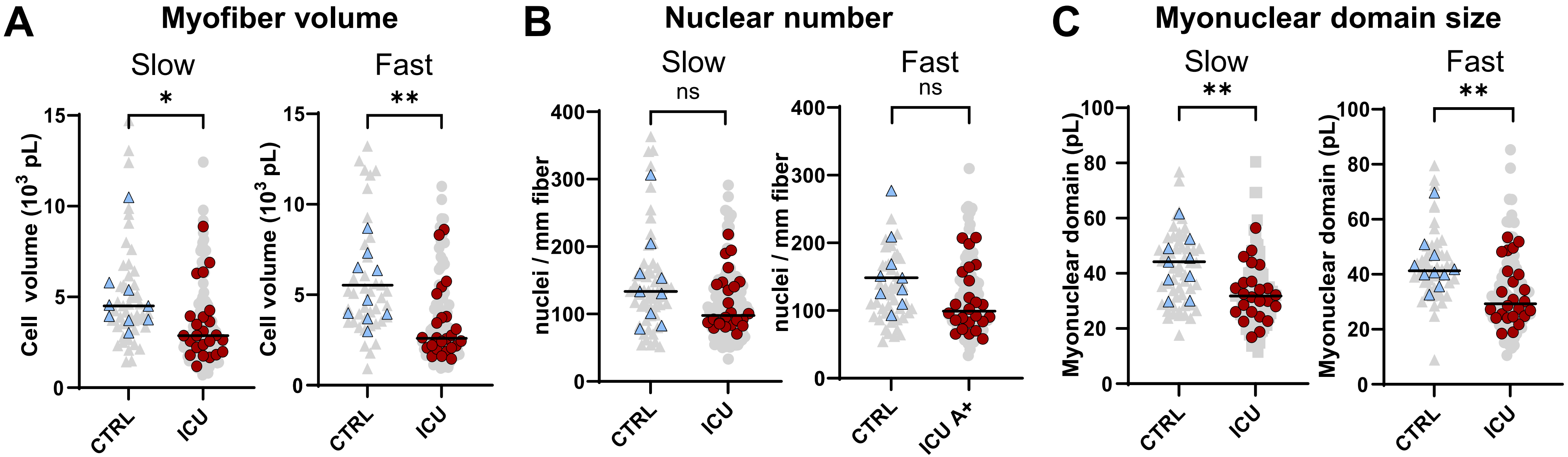
A
Myofiber volume
Slow
Fast
Cell volume ($10^3$ pL)
CTRL
ICU
B
Nuclear number
nuclei / mm fiber
ns
ICU A+
C
Myonuclear domain size
Myonuclear domain (pL)

**Figure S7**

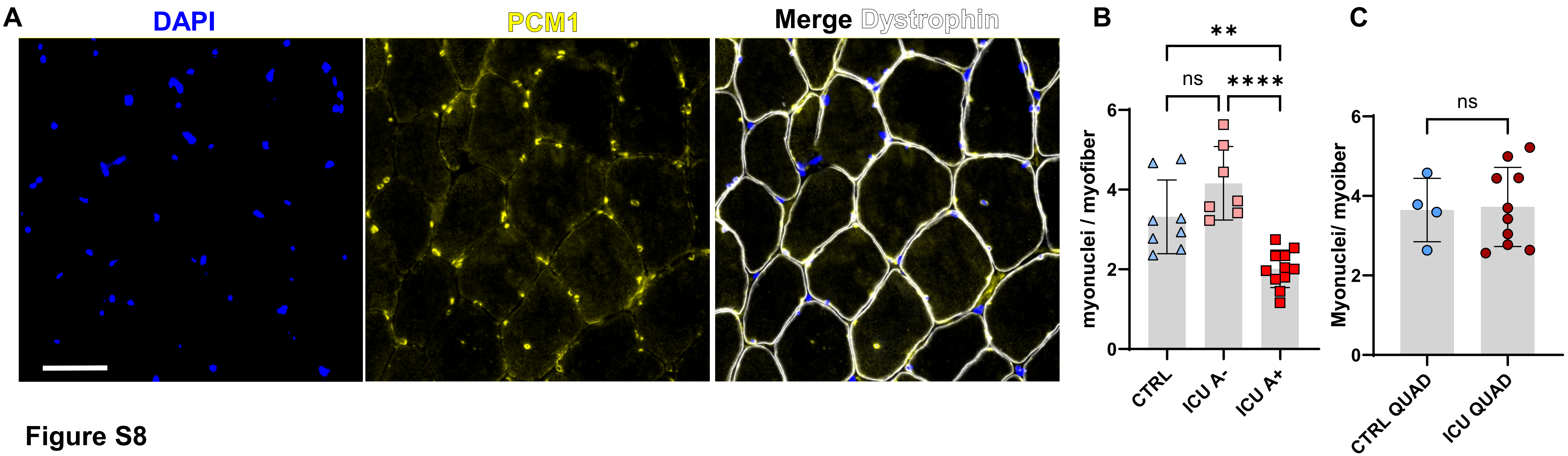
A
DAPI
PCM1
Merge Dystrophin
B
**
ns
****
6
4
2
0
myonuclei / myofiber
CTRL
ICU A-
ICU A+
C
ns
6
4
2
0
Myonuclei/ myoiber
CTRL QUAD
ICU QUAD

**Figure S8**

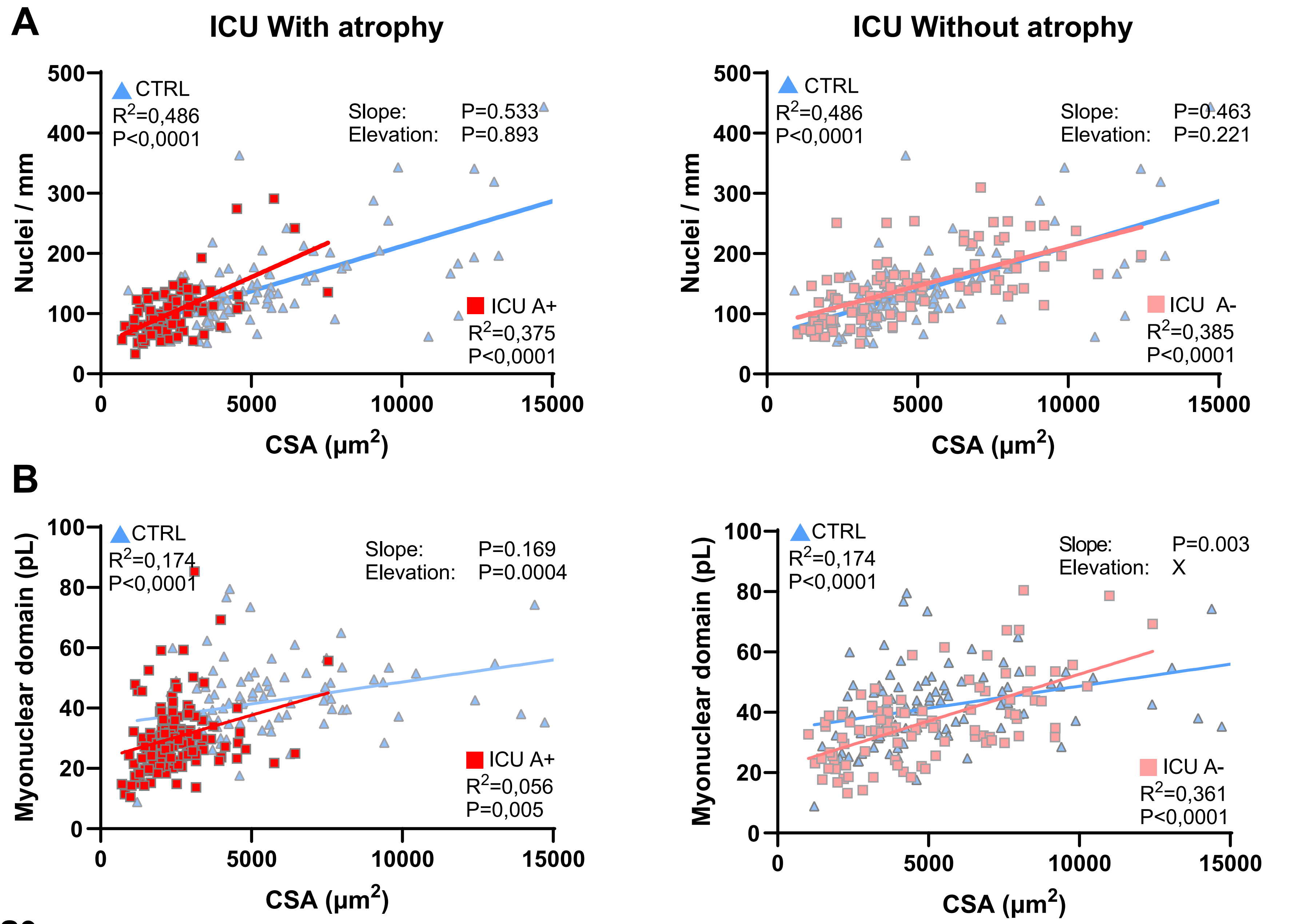

A
ICU With atrophy
ICU Without atrophy
CTRL
R2=0,486
P<0,0001
Slope: P=0.533
Elevation: P=0.893
ICU A+
R2=0,375
P<0,0001
Slope: P=0.463
Elevation: P=0.221
ICU A-
R2=0,385
P<0,0001
Nuclei / mm
CSA (µm2)
B
CTRL
R2=0,174
P<0,0001
Slope: P=0.169
Elevation: P=0.0004
ICU A+
R2=0,056
P=0,005
Slope: P=0.003
Elevation: X
ICU A-
R2=0,361
P<0,0001
Myonuclear domain (pL)
CSA (µm2)


Figure S9

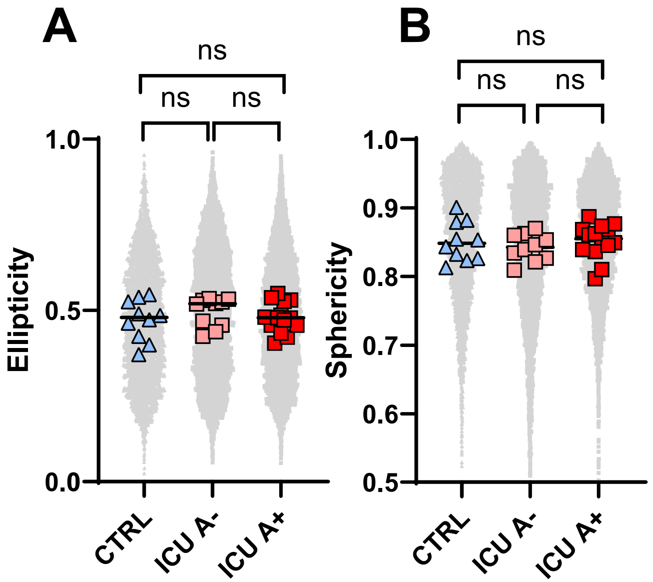
A
ns
ns
ns
1.0
0.5
0.0
Ellipticity
CTRL
ICU A-
ICU A+
B
ns
ns
ns
1.0
0.9
0.8
0.7
0.6
0.5
Sphericity
CTRL
ICU A-
ICU A+

**Figure S10**
